# A distinct serum metabolic signature outperforms C-reactive protein as a non-invasive marker for monitoring disease activity in Inflammatory Bowel Disease

**DOI:** 10.64898/2025.12.10.25341951

**Authors:** Lina Welz, Björn-Hergen Laabs, Danielle Harris, Sven Schuchardt, Florian Tran, Silvio Waschina, Norbert Frey, Andre Franke, Bram Verstockt, Séverine Vermeire, Philip Rosenstiel, Stefan Schreiber, Konrad Aden

## Abstract

**Background:** Achieving deep remission, such as e.g. histo-endoscopic remission, is a key objective in inflammatory bowel disease (IBD). However, this metric depends on invasive procedures that are impractical in routine care, whereas current non-invasive blood biomarkers demonstrate only limited correlation with disease activity.

**Objective:** We used serum metabolomics to identify blood-based signatures that comprehensively assess disease activity.

**Design:** Serum was collected from HC (n=195), UC (n=183; Kiel, discovery), UC (n=52; Leuven, validation) and CD (n=98; Kiel) cohorts alongside composite assessment of disease activity using endoscopic, clinical, biochemical, and histopathological criteria. Targeted metabolomics was performed with the Biocrates MxP Quant 500(XL) kit. Statistical analysis involved principal component analysis (PCA), linear mixed models (LMM), generalised estimating equation (GEE) and ML (logistic, LASSO regression).

**Results:** We identified numerous serum metabolites predominantly related to amino acid, bile acid and lipid metabolism that differed significantly between HC and inactive or active UC/CD. Among those, increased beta-alanine and Hex2Cer(d18:1/16:0) accompanied by diminished tryptophan and histidine distinguished active from inactive UC as well as HC from active IBD across several disease activity definitions. Combining those metabolites better estimated disease activity than C-reactive protein (CRP). Lastly, prior to initiation of advanced therapy, baseline metabolites and LASSO-selected combinations thereof exhibited limited ability to predict remission.

**Conclusion:** Serum metabolomics distinguished IBD from HC and inactive from active disease, with a four-metabolite panel outperforming CRP in comprehensively assessing disease activity. Despite restricted predictive performance, our findings underscore the value of serum metabolomics in elucidating IBD pathophysiology and improving disease monitoring.

**What is already known on this topic:** Serum metabolomics reveal systemic immunometabolic alterations in IBD, but prior studies lack composite disease activity measures and independent validation.

**What this study adds:** Cross-cohort analysis of comprehensively assessed disease activity revealed metabolic signatures that distinguish IBD from HC and inactive from active disease states, with superior discriminatory power compared to CRP. However, baseline metabolites exhibit limited ability to predict outcomes.

**How this study might affect research, practice or policy:** Our findings highlight both the potential and current constraints of serum metabolomics for clinical application, emphasizing the need for standardized, longitudinal, and multi-cohort studies to enable reproducible biomarker discovery in IBD.

## Introduction

IBD with its two main sub-entities CD and UC is a chronic inflammatory disorder of the intestine. Despite major scientific advances, its multifactorial aetiology remains incompletely understood. A disruption of the gut barrier coinciding with dysregulation of the complex interplay between environmental factors, genetic variants and host-microbial interactions is considered the central underlying pathophysiological principle triggering excessive immune responses [1,2]. Independent risk factors such as lifestyle and the exposome drive dysbiosis, aggravating immunological, microbial, and metabolic imbalances [3,4]. To date, therapies interfering with key immune pathways provide symptomatic relief but are not curative [5]. Consequently, only one-third of IBD patients achieve sustained long-term remission [6].

Several clinical endpoints have evolved to monitor IBD disease activity, including clinical, endoscopic, biochemical and histopathological remission. However, these endpoints have inherent limitations. While non-invasive biomarkers such as CRP and faecal calprotectin are widely use, their limited specificity and sensitivity restrict accurate disease monitoring and therapy guidance [7]. To overcome these shortcomings, endpoints are increasingly combined into composite measures capturing more holistic outcomes, such as histo-endoscopic remission or comprehensive disease control (CDC) [8,9]. Nevertheless, diagnostic procedures remain time-consuming, invasive, and burdensome for patients. Thus, there is a critical need for reliable, non-invasive markers that enable objective disease activity monitoring and predict disease courses.

Serum metabolomics enables to capture integrated fingerprints of host-microbiota interactions, immune activation, and metabolic dysregulation that may reflect IBD pathophysiology. Assessing metabolic alterations in IBD can therefore provide valuable insights into pathomechanisms and facilitate identification of novel biomarkers. Several studies have reported altered metabolic signatures in IBD, including disturbances in amino acid, bile acid, energy, and lipid metabolism [10,11]. However, prior work has been exploratory and cross-sectional with small sample sizes, often based on untargeted metabolomics and a limited range of metabolites. While associations between metabolic alterations and disease activity have been described, their relationship with clinical outcomes have not been systematically evaluated. Moreover, many prior studies did not distinguish between inactive and active disease states, lacked validation in independent cohorts, and relied on a single definition of disease activity, overlooking frequent discordance between clinical presentation and endoscopic inflammation. Consequently, the potential of metabolomic profiling as a monitoring or predictive tool in IBD remains underexplored.

By integrating targeted metabolomic profiling across cohorts and comprehensive definitions of disease activity, we aimed to identify alterations in key metabolic pathways that i) distinguish both HC from IBD as well as inactive from active disease states, and ii) predict subsequent remission of disease from baseline.

## Materials and methods

### Cohort design

Serum was sampled from HC (n=195; *Deutsche Zentrum für Herz-Kreislauf-Forschung* cohort [DZHK]) [12,13] (Table S1) and IBD patients. Longitudinal (n=92, Table S2) and cross-sectional UC cohorts (n=91, Table S3) from the University Hospital Schleswig-Holstein, Campus Kiel (UKSH), Germany, served as a combined discovery cohort. A longitudinal UC cohort (n=52, Table S4) from the University Hospital Leuven, Belgium, served as a validation cohort. In longitudinal studies, serum was sampled before (w0) and after (w14) initiating advanced therapy (Tables S2, S4). A limited number of samples were additionally collected at w2, w24, and w26 for UC cohorts. For CD (n=98, UKSH), longitudinal sampling was performed at w0, 2, 14, and 26.

Disease activity was monitored at each timepoint and based on the endoscopic Mayo score (eMayo), the histopathological Nancy score (only available for Kiel cohorts), clinical disease activity (PRO2), and serum CRP. For CD, measures included the Crohn’s Disease Activity Index (CDAI), the Simple Endoscopic Score for Crohn’s Disease (SES-CD), and CRP. Inactive UC was defined as eMayo≤1, PRO2≤1, Nancy≤1, or CRP≤5mg/l. Inactive CD was defined as CDAI≤220, SES-CD≤6 or CRP≤5mg/l. Endoscopic (eMayo≤1/SES-CD≤6) and clinical remission (PRO2≤1/CDAI≤220) were assessed in w14. Observations lacking disease activity metrics were omitted from respective sub-analysis. All metadata are provided in Table S1-5.

Studies at UKSH were approved by the Ethics Committee (Kiel University, D490/20, D489/20, A124/14, AZ156/03-2/13, EA1/300/15). Patients at University Hospital Leuven participated in the Institutional Review Board-approved IBD Biobank [B322201213950/S53684]. All patients provided written informed consent. Procedures followed national guidelines and the Declaration of Helsinki. Patients or the public were not involved in this research.

### Serum metabolomics

Serum was stored at -80°C until targeted metabolomics was performed using the Biocrates MxP Quant 500/XL kits (BIOCRATES Life Science AG, Innsbruck, Austria), which combine liquid chromatography-tandem mass spectrometry (LC-MS/MS) and flow injection analysis-tandem mass spectrometry (FIA-MS/MS) on an AB SCIEX QTRAP 5500 with electrospray ionization. Sample preparation and measurements followed the manufacturer’s instructions. Each analytical plate included calibrators and quality controls (QC) provided by the manufacturer. Data acquisition and peak integration were carried out in SCIEX Analyst/MultiQuant. Quantification, QC assessment, and concentration calculation were conducted in WebIDQ. Runs were accepted only if QC criteria met the manufacturer’s specifications. Metabolites common to both kits were merged to create a unified dataset. Values below the limit of quantitation and detection were considered missing and imputed as half of each metabolite’s minimum value, considering the respective thresholds of each kit. Metabolites present in ≥80% of samples were retained (80% rule [14]). Features with variance below the first quartile of variance distribution were removed to retain metabolites with higher biological relevance. Metabolites were log10 transformed prior to statistical analysis.

### Statistical analysis

To evaluate differences in overall metabolomic profiles between HC, inactive and active UC/CD, principal component analysis (PCA) was performed on metabolites residualised for age, gender, and repeated measures (and fasting for UC cohorts) using linear mixed models (LMM). Group differences were tested using permutational multivariate analysis of variance (PERMANOVA; 999 permutations, Euclidean distance). Pairwise PERMANOVA was conducted between all groups, with p-values adjusted using Benjamini-Hochberg (false discovery rate [FDR]) correction.

For LMM, age, gender, biologic class, and fasting (for UC Kiel cohorts, as cohort 2b was fasted; the other cohort was considered unfasted) were included as covariates. “Batch” was initially included as a covariate to account for using different kits and separate measurements in the Kiel cohorts but excluded from the final model due to collinearity with fasting and/or disease group. FDR-correction was applied with BH procedure. Unless stated otherwise, an FDR-adjusted p-value <0.1 was considered significant to balance discovery with type I error.

To estimate different measures of disease activity compared to CRP using the identified markers beta-alanine, Hex2Cer(d18:1/16:0, tryptophan, and histidine individually and in all possible combinations, generalised estimating equations (GEE) with exchangeable correlation structure accounting for within-subject correlations were employed. As the metabolic predictors were preselected, they were applied directly without additional model optimization. Separate estimation of area under the receiver operating characteristic curves (AUROC) was performed for the Kiel and Leuven cohort to avoid confounding from fasting-related effects in Kiel cohort 2b. Models comparing inactive vs. active UC included age, gender, and fasting (Kiel cohort; colinear with batch) as covariates. To compare HC vs. active IBD, we corrected for age and gender only, as fasting and batch were colinear with group status. “Biologic class” was omitted since, at w0, all patients had active disease, and none received biologics, resulting in perfect group estimation. Test-set AUC were derived from predicted probabilities obtained in out-of-bag data across 200 bootstrap iterations. During bootstrap resampling, training and test sets were defined at the subject level, preventing data leakage. For each iteration, the optimal probability threshold maximizing the Youden index was applied to the out-of-bag data to compute diagnostic metrics.

To identify baseline metabolic differences between w14 remitters and non-remitters, we assessed group differences using linear models including age, gender, and batch (Kiel cohort) as covariates. P-values were FDR-adjusted. To evaluate global metabolic patterns, baseline metabolites were residualised for age, gender, and batch (Kiel cohort) and analysed by PCA. Group separation was tested as described above.

Prediction of w14 remission based on baseline (w0) metabolites was performed in the longitudinal cohorts (Kiel, cohort 2a; Leuven, cohort 3). Least absolute shrinkage and selection operator (LASSO) models included all metabolites passing the 80% rule, without further prefiltering. Age and gender were incorporated as additional features. To predict endoscopic remission in w14 based on baseline metabolites, LASSO models were fitted using 10-fold cross-validation (CV) on the Kiel cohort and validated in the Leuven cohort. As 10-fold CV produced an instable model for clinical remission, a bootstrap-based approach (500 iterations) was used for this endpoint, with predictors selected in >50% of bootstrap samples considered stable. The predictive performance of *individual* metabolites for endoscopic and clinical remission was evaluated using logistic regression models trained on the Kiel cohort, adjusting for age and gender, and validated on the Leuven cohort. Only metabolites with concordant correlation directions with remission status across both cohorts were retained.

To investigate factors driving differences between the longitudinal Kiel and Leuven cohorts, PCA was performed including all available samples. Residualisation was carried out using LMM to adjust for age, gender, and batch.

Analyses were performed in R (version 4.4.1).

## Results

### Unique metabolic patterns differentiate inactive and active UC/CD from HC

We first evaluated overall changes of metabolite levels between disease groups (HC vs. inactive/active UC/CD) using principal component analysis (PCA). To prevent potential confounding of group separation by age, gender, fasting, or repeated measures, metabolites were residualised using linear mixed models (LMM). Disease activity was defined based on endoscopic and clinical features. Although group differences were statistically significant, small corresponding R² values suggested disease group explained little of the metabolomic variance.

We hypothesized that individual metabolites might still exhibit consistent and biologically meaningful differences between groups. We therefore assessed differential abundance at the single-metabolite level by applying LMM to compare HC to inactive and active UC/CD. Since assessment of disease activity cannot reliably be captured by single measures, we employed comprehensive definitions. Inactive UC was defined as eMayo≤1, PRO2≤1, Nancy≤1 (Kiel cohorts only), or CRP≤5 mg/l, and inactive CD as CDAI≤220, SES-CD≤6 or CRP≤5 mg/l. Based on these definitions, we identified numerous metabolites that were differentially abundant between HC and inactive or active UC/CD (Fig. S1). Each the top 50 metabolites ranked by FDR-adjusted p-value are displayed for every definition of disease activity for UC/CD (Fig. S2-4; complete lists are provided in Supplementary Tables 6-25).

We next focused on defining a unifying metabolic signature for both inactive and active disease as compared to HC. Metabolites were included if they were differentially abundant across all disease activity definitions and exhibited the same direction of change in each comparison. Since a UC validation cohort was available, only metabolites consistently identified in both cohorts were retained. Employing this approach, we found 21 metabolites to distinguish inactive UC vs. HC (Fig. S1, Fig. 2), whereas 59 metabolites differentiated active UC from HC (Fig. S1, Fig. 3). Differences between HC and inactive UC were dominated by alterations in amino and bile acid metabolism (glutamate, ornithine, taurine, beta-alanine, proline, glycochenodeoxycholic acid (GCDCA), creatinine) and various lipid classes, including di- and triacylglycerols (DGs, TGs), sphingolipids and cholesteryl esters (CEs) (Fig. 2, S2). Additionally, microbiota-associated metabolites (trimethylamine N-oxide [TMAO], 3-indolepropionic acid [3-IPA]), were reduced in inactive UC. Most alterations were retained in active disease vs. HC, while further changes involved metabolites associated with amino acid (aspartate, phenylalanine, tryptophan, homoarginine, methionine, cystine), purine (hypoxanthine), choline and microbial metabolism (indoxyl sulfate) (Fig. 3, S3). Several glycero-, phosphoand sphingolipids were elevated in active UC, whereas CEs were depleted.

**Fig. 1:**
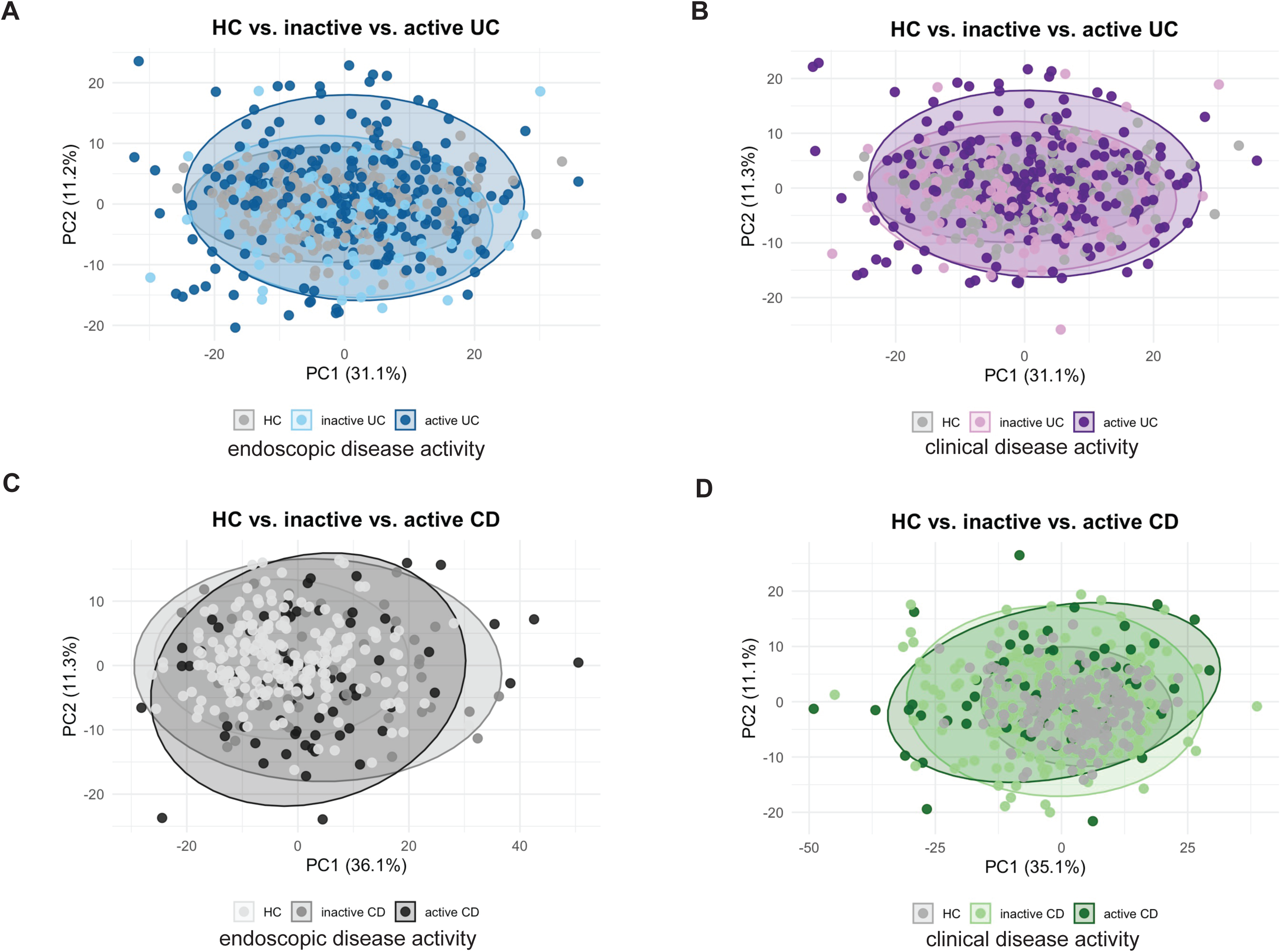
Unique metabolic patterns differentiate inactive IBD, active IBD and HC. Principal component analysis (PCA) was applied after adjusting baseline metabolites for age, gender, and repeated measures (and fasting in UC cohorts) using linear mixed models. Differences in overall metabolomic profiles among HC, inactive UC/CD, and active UC/CD were assessed using PERMANOVA (999 permutations, Euclidean distance). Data from all available UC cohorts were pooled for this analysis. Pairwise PERMANOVA comparisons between all groups were performed. P-values were corrected for multiple testing using the BH FDR method. (A) Disease activity based on eMayo. Inactive UC vs. HC: R² = 0.011, FDR p = 0.012; active UC vs. HC: R² = 0.018, FDR p = 0.003; inactive vs. inactive UC: R² = 0.007, FDR p = 0.034. (B) Disease activity based on PRO2. Inactive UC vs. HC: R² = 0.011, FDR p = 0.008; active UC vs. HC: R² = 0.019, FDR p = 0.001; inactive vs. active UC: R² = 0.006, FDR p = 0.051. (C) Disease activity based on SES-CD. Inactive CD vs. HC: R² = 0.03, FDR p = 0.0015; active CD vs. HC: R² = 0.03, FDR p = 0.0015; inactive vs. active CD: R² = 0.01, FDR p = 0.21. (D) Disease activity based on CDAI. Inactive CD vs. HC: R² = 0.018, FDR p = 0.003; active CD vs. HC: R² = 0.034, FDR p = 0.003; inactive vs. active CD: R² = 0.0051, FDR p = 0.2.

**Fig. 2:**
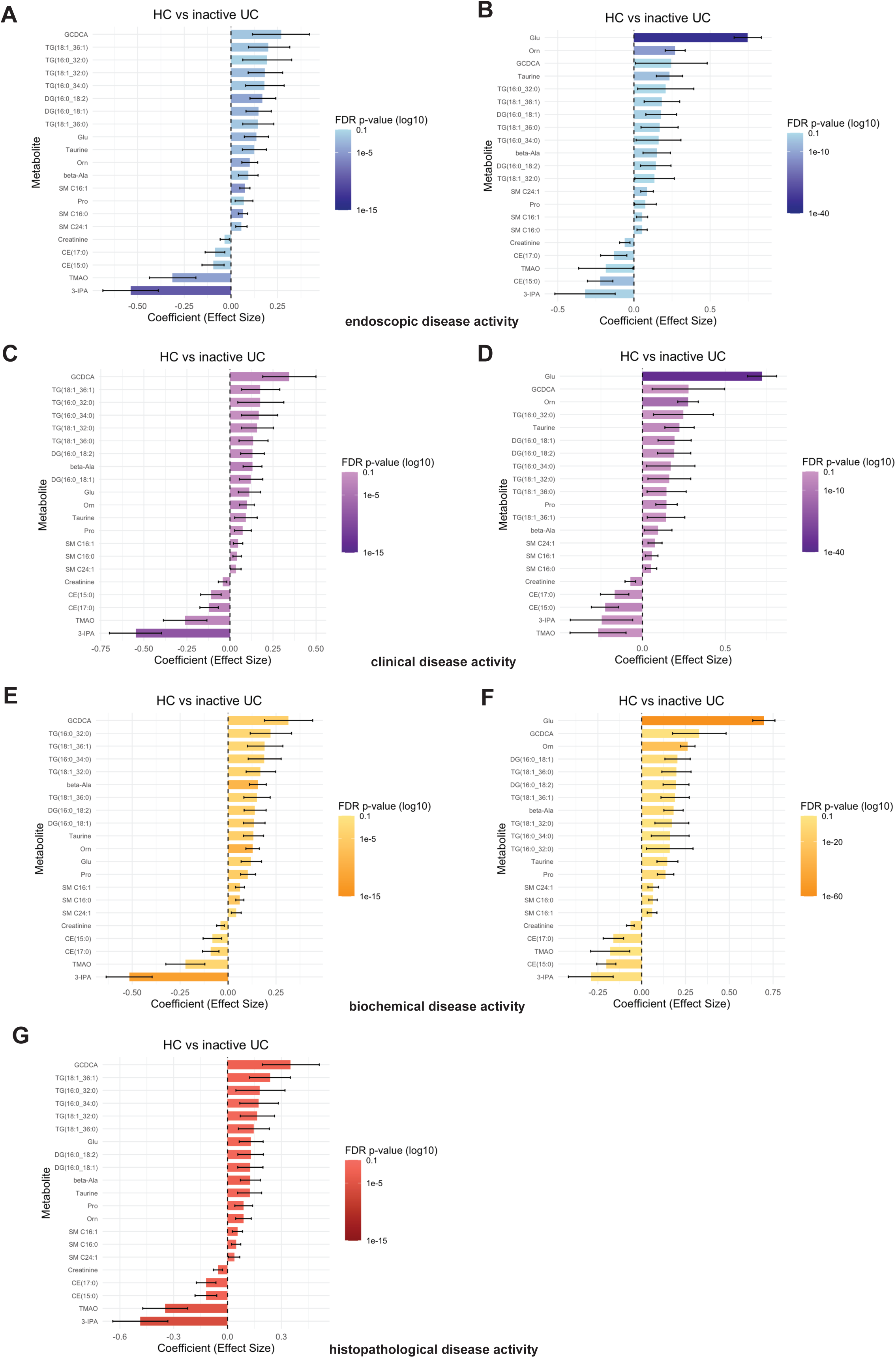
Unifying metabolic signature distinguishes inactive UC from HC. Linear mixed models (LMM) were applied to detect differentially abundant metabolites between HC and inactive UC. Definitions of disease activity were based on endoscopic (eMayo; A,B), clinical (PRO2; C,D), biochemical (CRP; E,F) and histopathological (Nancy; G) parameters in the Kiel (left) and Leuven cohort (right). LMM accounted for age, gender, biologic class, and fasting status (Kiel cohort). Multiple testing correction was performed using the BH FDR method (significance threshold <0.1). To identify a common metabolic signature, the displayed metabolites were selected based on consistent differential abundance and direction of change across all disease activity definitions and both cohorts. Glu: glutamate, Orn: ornithine, Pro: proline.

**Fig. 3:**
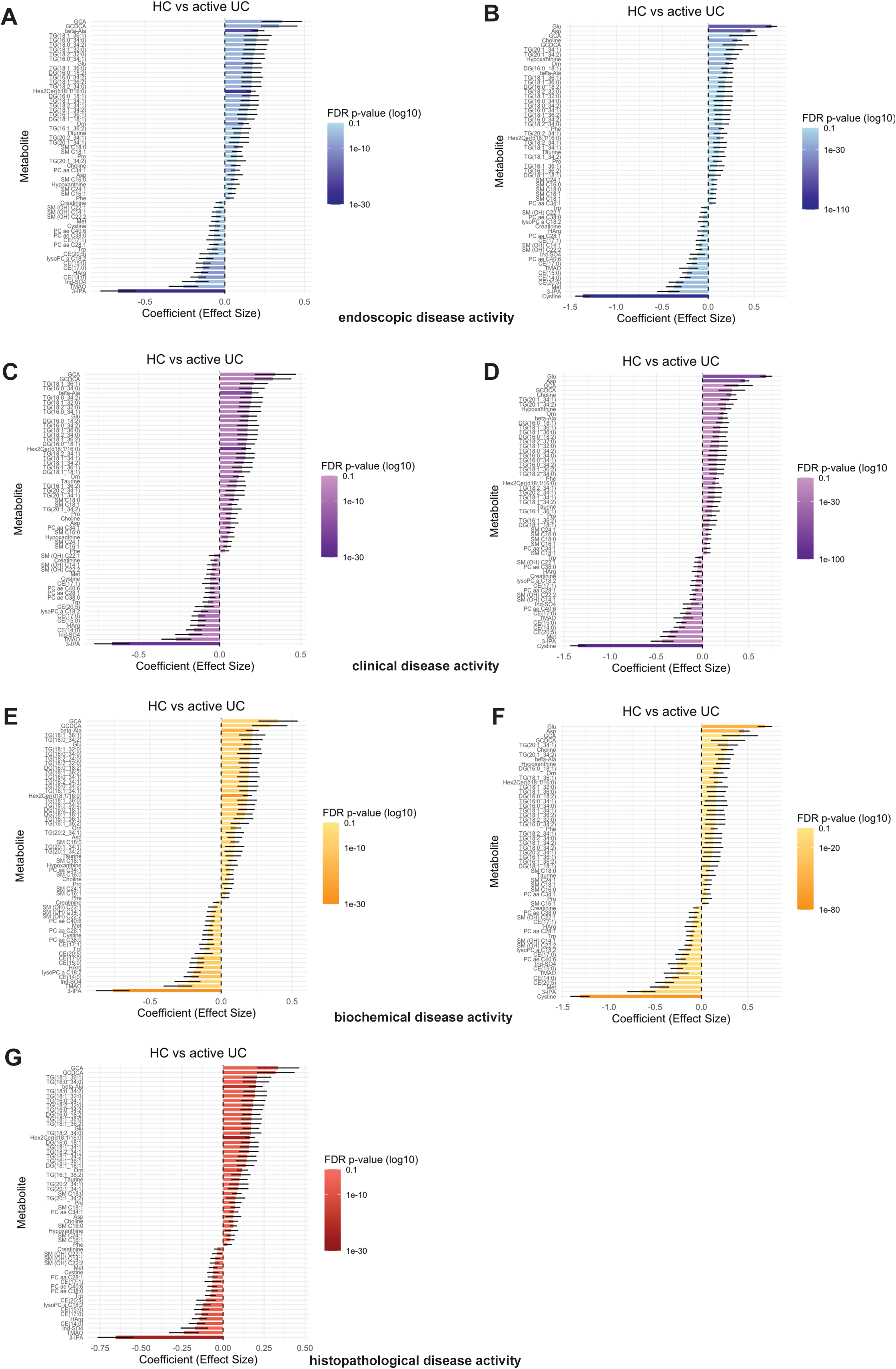
Common metabolic profile distinguishes active UC from HC. LMM were used to identify metabolites differing in abundance between HC and active UC. Disease activity was defined using endoscopic (eMayo; A,B), clinical (PRO2; C,D), biochemical (CRP; E,F), and histopathological (Nancy; G) criteria in the Kiel (left) and Leuven cohort (right). Models were adjusted for age, gender, biologic class, and fasting status (Kiel only). Multiple testing correction was applied using the BH FDR (FDR <0.1 significant). To derive a shared metabolic signature, the metabolites shown were selected for consistent differential abundance and direction of change across all disease activity definitions and both cohorts. Asp: aspartate, Glu: glutamate, Harg: homoarginine, Ind-SO4: indoxyl sulfate, Met: methionine, Orn: ornithine, Phe: phenylalanine, Pro: proline, Trp: tryptophan.

A large number of metabolites discriminated inactive (n=243) and active CD (n=267) from HC based on a composite definition of disease activity (endoscopic, histopathological, biochemical), of which each the top 50 metabolites ranked by FDR-p value are displayed (Fig. S1, S5). The numbers of differentially abundant metabolites are, however, likely overestimated without a CD validation cohort. We found active CD was characterized by a widespread upregulation of lipids (mainly TGs and sphingomyelins [SMs]) and bile acids, accompanied by downregulation of CEs, hydroxylated SMs, microbial indole derivatives (3-IPA, 3-indoleacetic acid), and phospholipids. While increased levels of energy- and nitrogen-related amino acids (glutamate, aspartate, ornithine, beta-alanine, proline, taurine) where observed in active CD, tryptophan, histidine, methionine, threonine, tyrosine, valine, citrulline, homoarginine, and cystine were reduced.

Several metabolites were consistently differentially abundant between inactive UC/CD and HC: 3-IPA, beta-alanine, CE(15:0), CE(17:0), creatinine, DG(16:0/18:1), DG(16:0/18:2), TG(16:0/32:0), TG(16:0/34:0), TG(18:1/32:0), TG(18:1/36:0), TG(18:1/36:1), GCDCA, glutamate, ornithine, proline, SM(16:0), taurine, and TMAO. The following metabolites distinguished both active UC/CD from HC: 3-IPA, aspartate, beta-alanine, CE(14:0), CE(15:0), CE(17:0), CE(17:1), CE(20:5,) choline, creatinine, cystine, DG(16:0/18:1), DG(16:0/18:2), DG(18:1/18:1), glycocholic acid, GCDCA, glutamate, homoarginine, Hex2Cer(d18:1/16:0), hypoxanthine, lysophosphatidylcholine(C18:2), methionine, ornithine, diacyl phosphatidylcholines (PCaa)(C28:1), PCaa(C34:1), alkyl-acyl phosphatidylcholine (PCae)(C38:0), PCae(C40:6), phenylalanine, proline, several (hydroxylated) SMs, TGs, taurine and tryptophan. These overlapping metabolic signatures indicate both IBD entities and activity states share a core set of metabolic disturbance compared to HC. The broader range of metabolites distinguishing active disease suggests additional shifts associated with increased inflammation.

Overall, metabolic alterations identified in IBD affect bile and amino acid metabolism and complex lipid homeostasis, reflective of heightened energy and nitrogen turnover, oxidative stress, membrane remodelling, and disrupted host-microbiome interaction.

### A distinct metabolic signature distinguishes inactive and active UC

We next asked whether overlapping metabolic signatures are likewise found when comparing inactive to active IBD. In contrast to the previous analysis of HC versus IBD, comparing inactive and active disease states yielded only a small number of differentially abundant metabolites, indicating high phenotypic similarity (Fig. 4). To nonetheless identify unifying metabolic signatures for all definitions of disease activity in our two UC cohorts, we adopted a modified strategy: we first applied LMM to the Kiel cohort to select metabolites with an FDR-adjusted p-value <0.5 and subsequently validated these candidate metabolites in the Leuven cohort. Following this approach, we selected four metabolites reflective of a shared metabolic signal across disease activity definitions and cohorts, although some were absent in individual analyses: increased levels of beta-alanine and dihexosylceramide (Hex2Cer)(d18:1/16:0), together with diminished abundances of tryptophan and histidine, indicated active compared to inactive UC (Fig. 4; metabolic abundances Fig. S7).

**Fig. 4:**
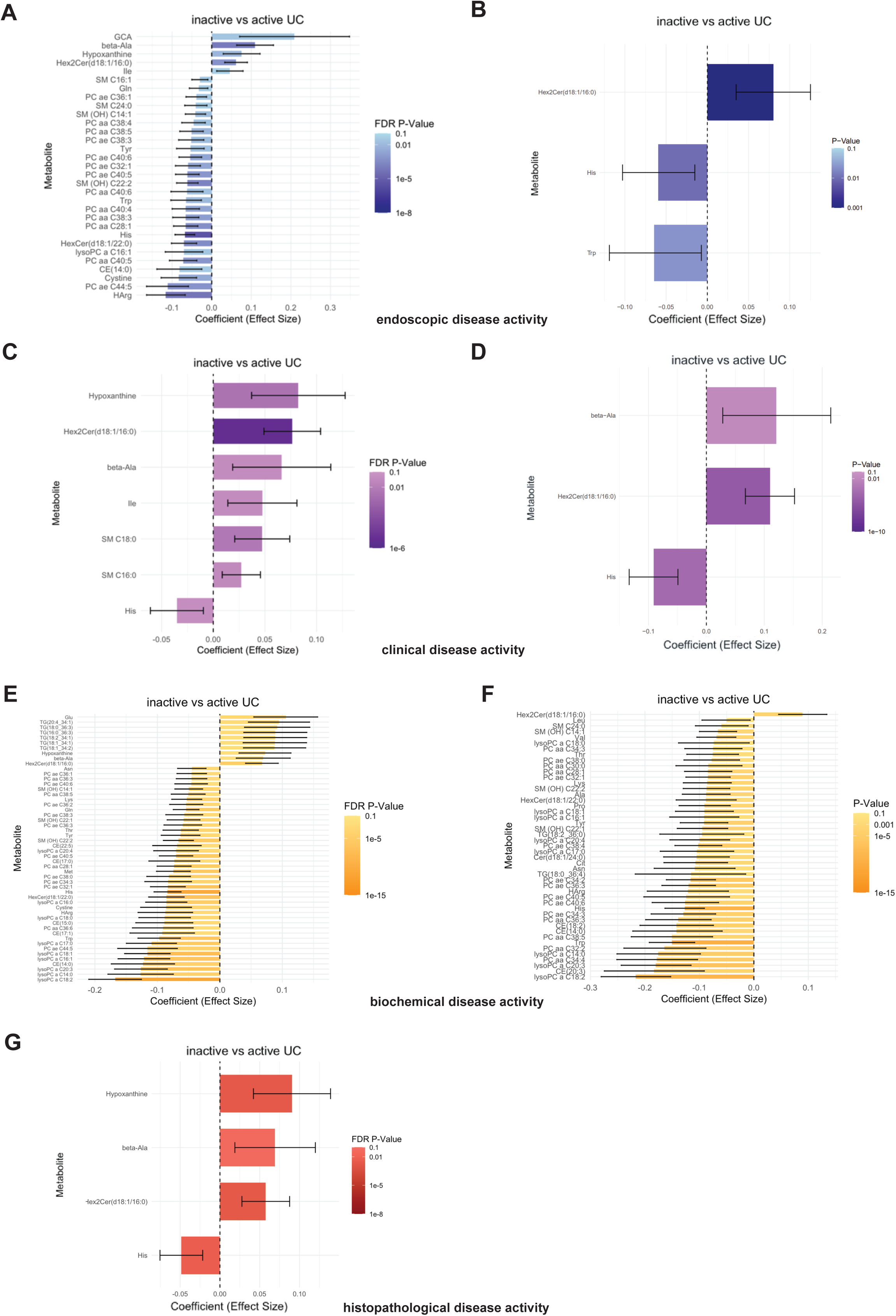
Shared serum metabolites discriminate inactive from active UC. LMM were applied to detect differentially abundant metabolites between inactive and active UC. Definitions of disease activity were based on endoscopic (eMayo; A,B), clinical (PRO2; C,D), biochemical (CRP; E,F), and histopathological (Nancy; G) parameters in the Kiel (left) and Leuven cohort (right). LMM accounted for age, gender, biologic class, and fasting status (Kiel cohort). To derive a common signature, LMM were first applied to the Kiel cohort to identify candidate metabolites using an FDR-adjusted p-value cutoff of 0.5 and subsequently validated in the Leuven cohort. Multiple testing correction was performed using the BH FDR method (significance threshold <0.1).

However, when comparing inactive to active CD, no unifying metabolic signature was detectable, although glutamate was upregulated and lysoPCaC20:3 downregulated across two disease activity definitions (Fig. S6). Elevated Hex2Cer(d18:1/16:0) and reduced histidine also characterized active endoscopic CD, mirroring patterns observed in active UC.

### Serum metabolic signature identifies active IBD and outperforms CRP in comprehensively assessing disease activity

Building on our identified serum metabolic signature of active UC, we evaluated its performance in discriminating endoscopic, clinical, and – for the Kiel cohorts – histological disease activity in patients with inactive and active UC in comparison to CRP. Testing all possible combinations of beta-alanine, Hex2Cer(d18:1/16:0), tryptophan, histidine, and CRP, we observed that combinations of these metabolites – either including or excluding CRP – consistently outperformed CRP as a single marker in estimating each disease activity definition in the Kiel and Leuven cohort (Fig. 5A-E). Integrating endoscopic, clinical, histological (Kiel only), and biochemical measures of disease activity to approximate CDC, we again observed that combinations of the selected metabolic panel were superior to CRP alone (Fig. 5F,G; sensitivity and specificity are supplied in the figure legends).

**Fig. 5:**
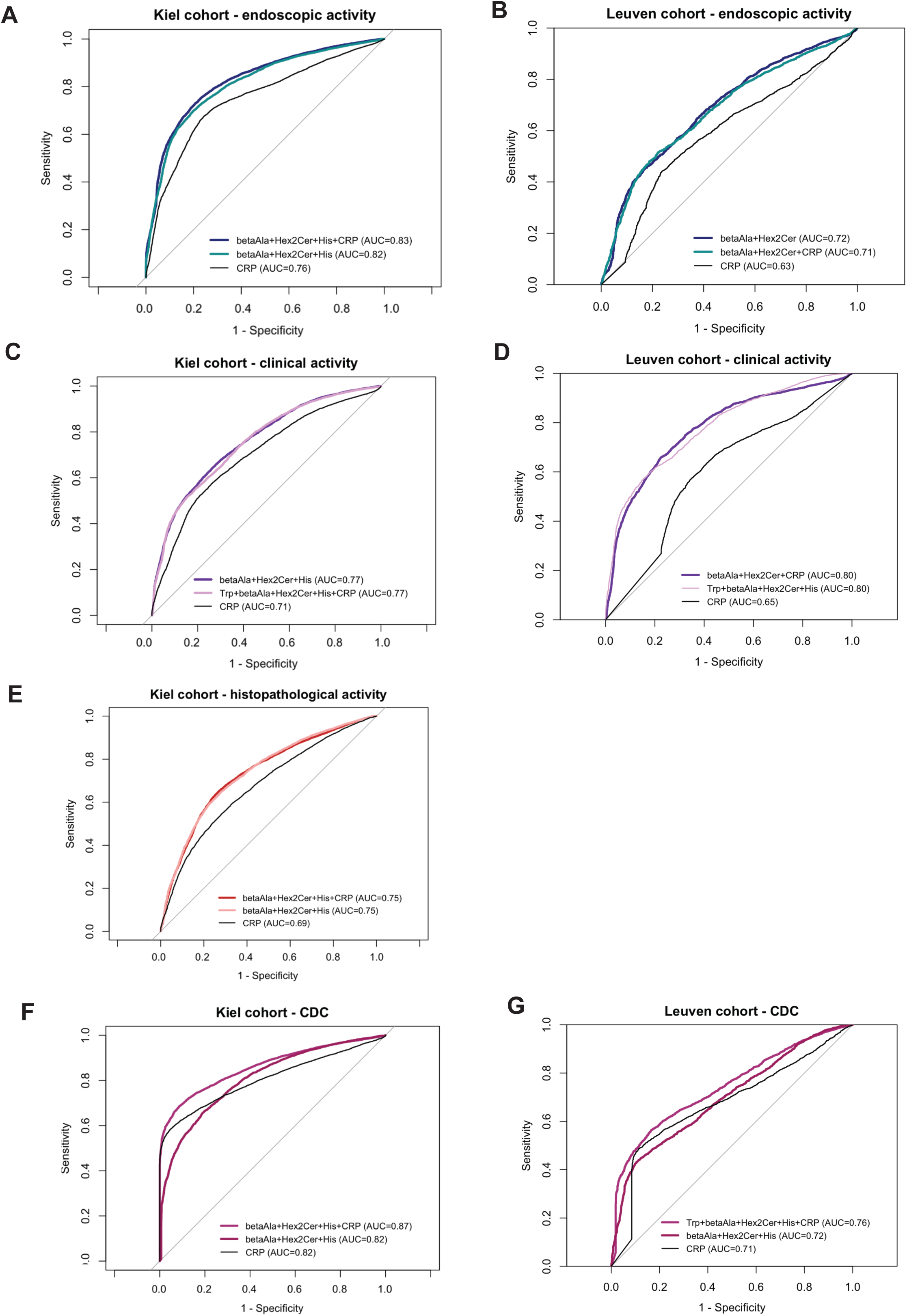
Serum metabolites outperform CRP in comprehensively estimating disease activity. To estimate different definitions of disease activity, the identified serum markers beta-alanine, Hex2Cer(d18:1/16:0), tryptophan, histidine, as well as CRP were evaluated individually and in all possible combinations using GEE. AUCs were estimated separately for the Kiel (left) and Leuven cohort (right). Models comparing the indicated measures of inactive vs. active UC included age, gender, and, for the Kiel cohorts, fasting as covariates. Both the highest AUC values for metabolite combinations with and without CRP are displayed in comparison to the AUC for CRP alone. Sensitivity and specificity determined with the Youden index of the models with the highest AUC as compared to CRP were as follows: (A) 0.76/0.77 vs. 0.69/0.74; (B) 0.63/0.61 vs. 0.56/0.62; (C) 0.68/0.68 vs. 0.61/0.69; (D) 0.71/0.73 vs. 0.64/0.58; (E) 0.71/0.67 vs. 0.63/0.61; (F) 0.75/0.83 vs. 0.65/0.85; (G) 0.67/0.7 vs. 0.58/0.77.

Given that elevated beta-alanine and Hex2Cer(d18:1/16:0) levels as well as decreased tryptophan concentrations were also found when comparing active UC with HC, and that active CD versus HC exhibited a similar pattern (increased beta-alanine and Hex2Cer(d18:1/16:0) together with reduced tryptophan and histidine levels), we further evaluated whether the metabolite panel could furthermore distinguish active IBD from HC. Indeed, their combinations showed strong discrimination of active IBD vs. HC (Fig. S8,S9). CRP values were not available for HC.

Overall, we find that combining beta-alanine, Hex2Cer(d18:1/16:0), tryptophan, and histidine robustly identifies active IBD based on composite measures of disease activity and outperforms CRP.

### Baseline serum metabolomics have limited capacity to predict remission in UC

We lastly tested the ability of baseline serum metabolites to predict endoscopic and clinical remission in w14 in UC patients initiating biologic treatment.

To examine general baseline differences in the metabolome (w0) stratified by endoscopic or clinical remission (R) or non-remission (NR) in w14, we applied PCA but did not observe major distinctions between groups (Fig. 6). Linear models revealed that hypoxanthine was the only baseline metabolite significantly differing between endoscopic R and NR in the Kiel cohort (data not shown).

**Fig. 6:**
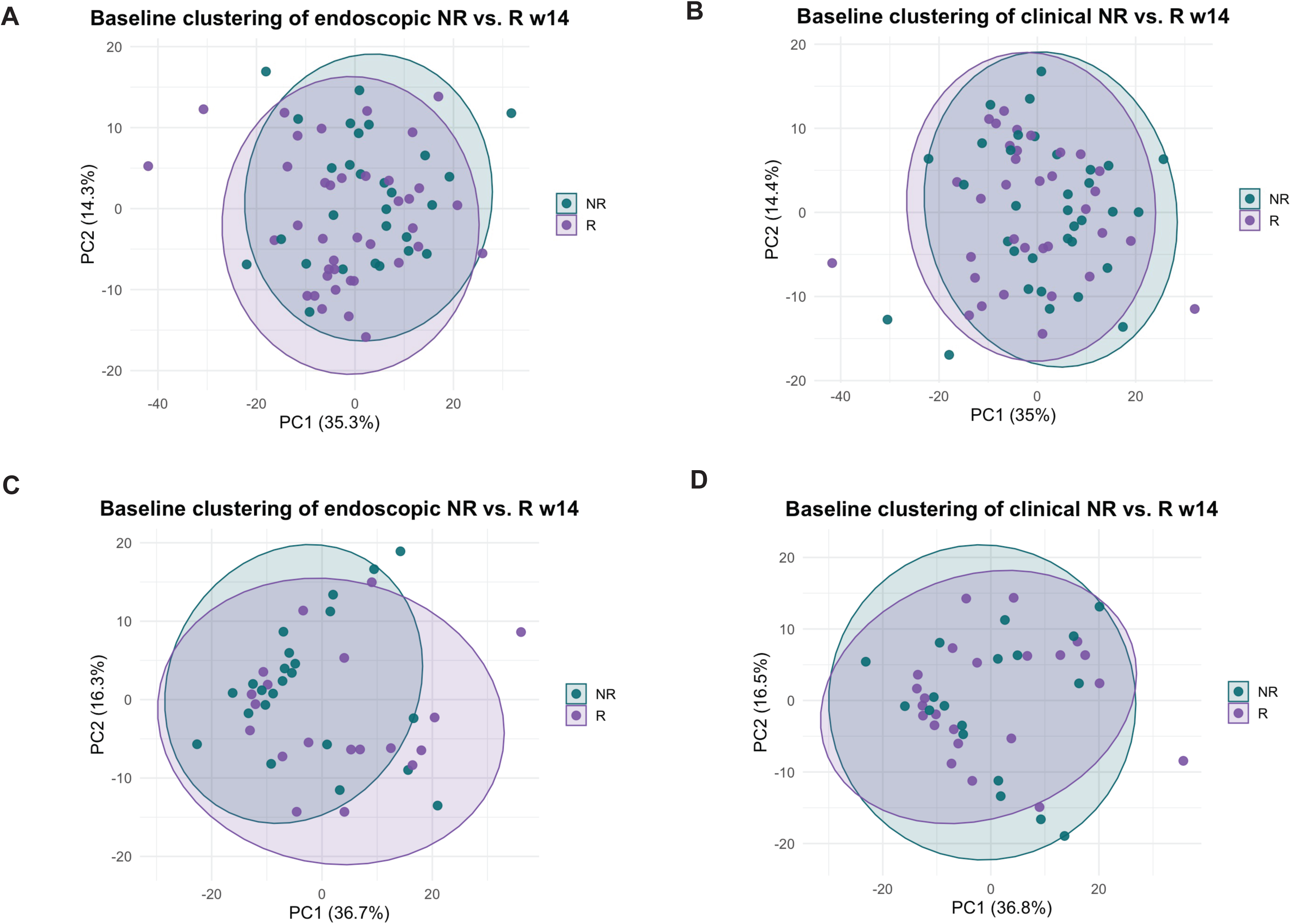
Baseline metabolome stratified by remission status in w14. For evaluation of global baseline metabolic patterns between remitters (R) and nonremitters (NR) at w14, baseline metabolite data were residualised for age, gender, and batch (Kiel cohort) and analysed by PCA. Group separation was tested using PERMANOVA with 999 permutations and Euclidean distance. PCA plots show clustering of baseline metabolic patterns stratified on endoscopic (A, R² = 0.02, p = 0.22; C, R² = 0.05, p = 0.086) or clinical (B, R² = 0.02, p = 0.2; D, R² = 0.01, p = 0.96) remission in w14 for the Kiel (left) and Leuven cohort (right).

We thus employed LASSO regression to select a combination of baseline metabolites best predicting w14 remission status. Whereas a robust cross-validated AUC for predicting endoscopic remission was achieved in the Kiel cohort (0.8), the model predicting clinical remission performed remarkably worse (0.62), with the discriminative ability of both models further declining in the validation cohort (AUC 0.63/0.57) (Fig. 7A,B).

**Fig. 7:**
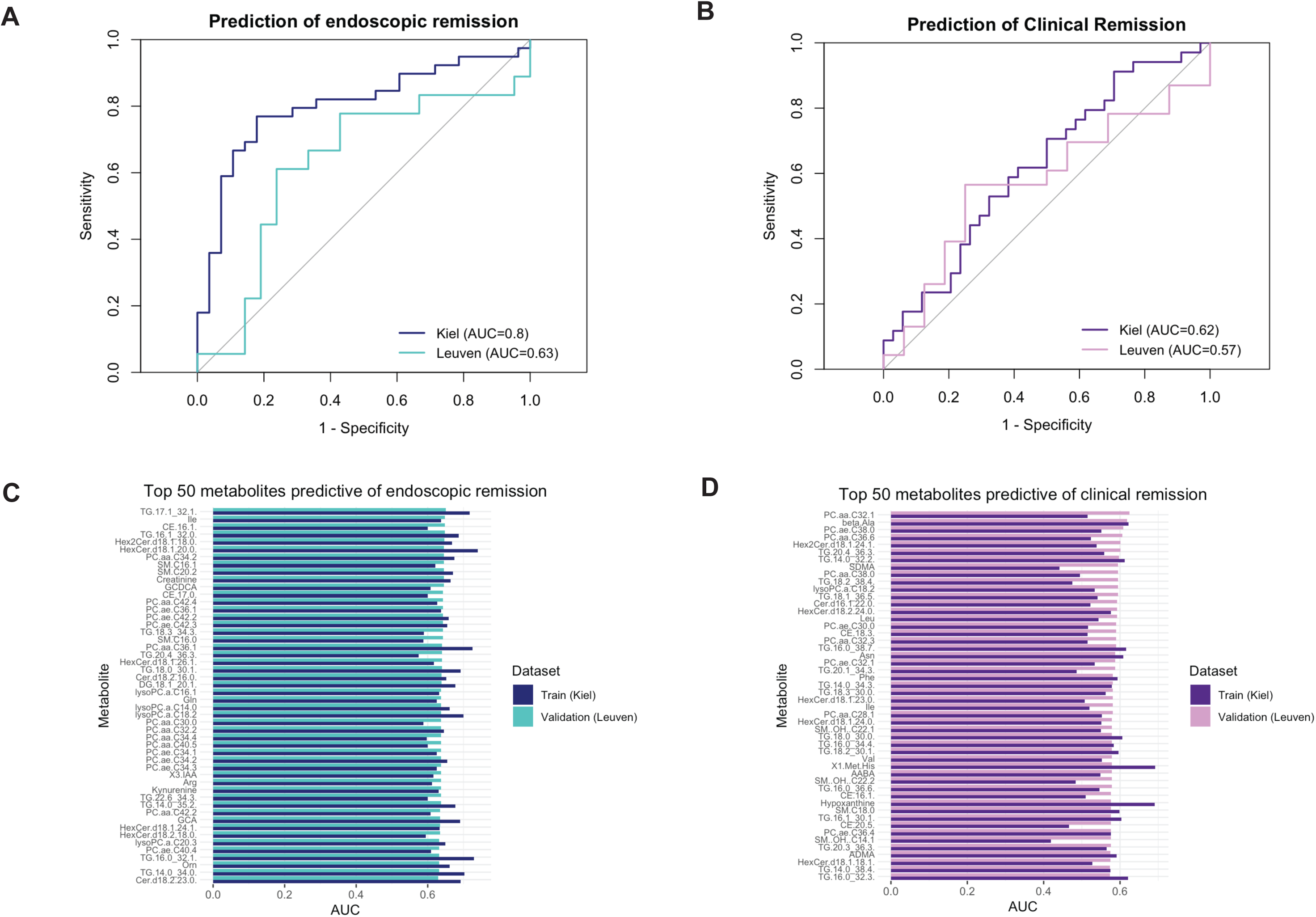
Baseline serum metabolites predict remission in UC. Prediction of remission in w14 based on baseline (w0) metabolite levels was performed exclusively in longitudinal cohorts. (A,B) LASSO models included all metabolites passing the 80% rule, with age and gender incorporated as additional features. Models were fitted using 10-fold CV or bootstrapping on the Kiel cohort and validated on the independent Leuven cohort to predict endoscopic (A) and clinical (B) remission in w14. Selected predictors included (A) tyrosine, citrulline, PCaaC40:3, Hex2Cer(d18:1/14:0), Hex2Cer(d18:1/20:0), Hex3Cer(d18:1/24:1), hypoxanthine, H1 (hexoses, including glucose), TG16:0/37:3., 1-methylhistdine, α-aminobutyric acid, cysteine, serotonin, Cer(d18:1/26:1), PCaeC38:1, TG(17:2/34:2), and age and (B) carnitine, arginine, lysine, citrulline, Cer(d18:0/24:0), Hex3Cer(d18:1/18:0), Hex3Cer(d18:1/24:1), HexCer(d18:2/20:0), hypoxanthine, 1-methylhistidine, TG(18:3/38:5), asymmetric dimethylarginine, CRP, and gender. (C,D) The predictive performance of individual metabolites for endoscopic and clinical remission was evaluated using logistic regression models trained with 10-fold CV on the Kiel cohort, adjusting for age and gender, and externally validated on the Leuven cohort. Training AUCs were received using held-out folds during CV.

Since the reduced performance of the multi-metabolite models likely reflected cohort-specific regulation of individual metabolites, we assessed the prognostic value of each baseline metabolite independently using logistic regression trained on the Kiel cohort and validated on the Leuven cohort. Following this approach yielded cross-validated training AUCs of up to 0.8 and 0.75, with a maximum AUC of 0.65 for endoscopic and 0.63 for clinical remission in the validation cohort (Fig. 7C,D). Individual metabolites demonstrated superior predictive performance compared to CRP (validation AUC: endoscopic 0.6/clinical remission 0.57; not shown). However, no metabolite was significantly associated with remission status in the validation cohort. Together with low AUCs, this indicated limited generalisability of predictive models to external cohorts.

To explore potential causes of the reduced external performance, we conducted another PCA including baseline and w14 data. A marked within-patient metabolic shift between w0 and w14 was evident in the Leuven cohort but absent in the Kiel cohort, indicating individual variability in metabolic trajectories toward remission likely impairs cross-cohort prediction (Fig. S10). Importantly, proportions of active disease at baseline and patients achieving remission were comparable across cohorts.

Overall, although substantial heterogeneity limited the ability of serum metabolomics to predict disease trajectories in IBD, we identified a metabolic serum signature that outperformed the routine non-invasive biomarker CRP in comprehensively capturing disease activity. Thus, despite the variability inherent to IBD, metabolomic profiles provide a sensitive snapshot of inflammatory activity to guide disease monitoring.

## Discussion

Serum metabolomics has emerged as a powerful tool to characterize IBD. Here, we integrated targeted serum metabolomic profiling across independent cohorts using comprehensive clinical, endoscopic, biochemical, and histopathological criteria. To our knowledge, a similar approach has not been undertaken before.

In line with previous reports, we observed numerous differentially abundant metabolites between HC and UC/CD. Yet, no distinct clustering was apparent in PCA, suggesting that disease-related changes, although widespread, are subtle compared with biological variability among individuals. We thus aimed to capture a robust metabolic signal reflecting association of individual metabolites with composite definitions of disease activity in two independent cohorts. Metabolic alterations distinguishing HC from inactive UC were dominated by changes in amino and bile acid metabolism and increased lipids. Most alterations persisted in active UC, with additionally elevated glycerolipids, phospholipids and sphingolipids, alongside depletion of CEs. Active CD exhibited broad shifts with increased lipids and bile acids, reduced CEs, microbial indole metabolites, and altered amino acid metabolism. Substantial overlap of metabolites differentially abundant in CD and UC compared to HC reflected convergent patterns of metabolic dysregulation independent of disease entity, with a wider range of metabolites distinguishing active inflammation. These alterations align with inflammation-driven immunometabolic reprogramming in several biological compartments. Prior studies similarly reported perturbations in amino and bile acid, lipid, and energy metabolism in IBD patients’ serum [10,11,15]. Overall, these reflect inflammation-induced perturbations in redox balance, energy utilisation, and barrier function, as indicated, for instance, by reduction of antioxidative histidine and methionine or accumulation of lipid-related metabolites, consistent with lipolysis and membrane remodelling [16,17].

Shifts in amino acids and related metabolites represented a predominant pattern. Whereas previously described alterations of branched-chain amino acids were not evident, glutamate, aspartate, phenylalanine, proline, ornithine, beta-alanine and taurine were elevated in active IBD, suggesting enhanced TCA and urea cycle activity as well as antioxidant responses [11]. Depleted tryptophan, histidine, and methionine levels in active IBD were consistent with our previous analysis across systemic chronic inflammatory disorders, which revealed reduction of predominantly essential amino acids, indicating increased protein synthesis [12]. Whereas directionalities of amino acid alterations were inconsistent across studies, presumably reflecting methodological and cohort-related heterogeneity, our integrative approach allows contextualization of prior findings within a comprehensive framework of disease activity.

Consistent with our findings, diminished tryptophan and histidine stood out robust across previous reports [18–20]. We and others have shown serum tryptophan depletion to result from enhanced degradation along the kynurenine pathway (KP) in chronic inflammation, leading to elevated serum kynurenine, which we observed in active IBD in the Kiel cohorts [12,21,22]. KP activation was paralleled by reduced microbial tryptophan catabolites 3-IPA and indoxyl sulfate. We have recently shown that IPA provides protection by enhancing mitochondrial respiration [23].

Prior studies suggested histidine is anti-inflammatory and predictive of relapse in UC [24–27]. Here, baseline histidine was not predictive of remission status. Interestingly, low histidine was also observed in rheumatoid arthritis and chronic kidney disease [28,29].

Associations of IBD and beta-alanine are rarely described [30,31]. Beta-alanine is generated, among other pathways, through pyrimidine or vitamin B5 degradation, and is both a product and substrate for histidine-containing dipeptides (anserine, carnosine). Although elevated beta-alanine and reduced histidine, accompanied by decreased creatinine in active IBD, do not allow a definitive conclusion, they point toward muscle protein catabolism.

Limited literature is available on Hex2Cer(d18:1/16:0). However, recent studies correlated long-chain ceramides with both IBD diagnosis and severity [32,33].

The four-metabolite marker set discriminated well between active and inactive UC based on comprehensive disease activity measures and exceeded CRP in performance; it also distinguished HC from active IBD. In line with previous reports, we observed higher specificity than sensitivity for CRP [34]. Whereas true external validation was not feasible due to the need to control for fasting status – the Kiel cohort included a fasted sub cohort, while this information was lacking for the Leuven cohort – the primary objective remained comparison with CRP, which served as both a validated biomarker and a benchmark, given its inclusion in our composite disease activity assessment. Notably, in terms of clinical applicability, beta-alanine, histidine, and tryptophan, which are measurable by standard HPLC in contrast to Hex2Cer(d18:1/16:0), were sufficient to estimate disease activity and still outperformed CRP. However, the panel did not predict w14 remission status at baseline (not shown).

Whereas serum metabolites distinguished IBD disease states from each other and from HC, they exhibited limited potential to predict w14 remission status. Although training AUCs generated via internal cross-validation were moderate, external validation demonstrated poor generalizability. To date, very few IBD studies included a true independent validation cohort for predictive modelling, with the majority relying on internal cross-validation or similar train-test partitioning within the same cohort – a practice that, as illustrated here, likely leads to substantial overestimation [35]. One study utilising a proper external validation cohort consistent with our findings reported that neither microbial nor metabolomic features reliably predict response to advanced IBD therapies [36]. Another study, which validated models predictive of later IBD onset across three large national serum metabolomics biobanks, demonstrated AUCs between 0.62 and 0.76, highlighting that limitations in predictive capacity are not simply attributable to small cohort size [37].

The lacking predictive power may be due to differences in disease history, host-microbiome interactions, diet, environment, or medication, contributing to substantial individual heterogeneity. Additionally, we observed pronounced metabolic changes over the treatment course in the Leuven cohort, whereas such changes were scarcely detectable in the Kiel cohort. Similarly, also CRP did not predict remission at baseline. Thus, not only the direction but also the magnitude of metabolic alterations may affect external transferability.

Some limitations should be acknowledged. First, faecal calprotectin represents a more accurate marker of IBD disease activity than CRP; however, this parameter was largely unavailable. Data coverage for disease extent and steroid use was insufficient as well. Second, histopathological information was lacking for the Leuven cohort. Lastly, a validation cohort for CD was unavailable, limiting the most relevant findings to UC.

In conclusion, this study integrates targeted serum metabolomics with a composite assessment of disease activity across independent cohorts, representing a novel approach to comprehensively characterize disease activity in IBD that outperforms CRP and reflects underlying immunometabolic inflammation. Although the predictive capacity of baseline metabolites was limited, we identified a non-invasive, four-metabolite panel that robustly captures intestinal inflammation and demonstrates potential as a clinically accessible biomarker of disease activity.

## Author Contributions

L.W., S.V., K.A., P.R., S.Schr., designed the study.

L.W., D.M.M.H., B.-H.L., B.V., S.Sch., F.T., S.W. performed experiments and analysed the data.

L.W., K.A. planned the project and supervised the experiments.

L.W., K.A. wrote the initial manuscript.

K.A., B.-H.L., S.V., P.R., S.Schr. edited the manuscript

## Data availability

Serum metabolomics data are accessible via the Zenodo repository using the following link: https://doi.org/10.5281/zenodo.17855140.

## Abbreviations

3-IPA: 3-indolepropionic acid
AUC: area under the curve
CD: Crohn’s disease
CE: cholesteryl ester
CRP: C-reactive protein
DG: diacylglycerol
GCDCA: glycochenodeoxycholic acid
GEE: generalised estimating equation
HC: healthy controls
IBD: inflammatory bowel disease
KP: kynurenine pathway
LMM: linear mixed models
NR: non-remission / non-remitters
PCA: principal component analysis
PCaa: diacyl phosphatidylcholines
PCae: alkyl-acyl phosphatidylcholine
R: remission / remitters
SM: sphingomyelin
TG: triacylglycerol
TMAO: trimethylamine N-oxide
UC: ulcerative colitis

---

**Supplementary Table 1.** Cohort 1: DZHK cohort, HC.

|  |  |
| --- | --- |
| Number of patients (%) | 195 |
| Number of observations (%) | 195 |
| Gender (%) |  |
| Male | 108 (55.4) |
| Female | 87 (44.6) |
| Age (mean (SD)) | 46.82 (8.3) |
| BMI (mean (SD)) | 24.74 (3.8) |

**Supplementary Table 2.** Cohort 2a: Kiel cohort, longitudinal UC discovery cohort.

|  |  |
| --- | --- |
| Number of patients | 92 |
| Number of observations | 166 |
| Gender (%) |  |
| Male | 46 (50) |
| Female | 46 (50) |
| Age at baseline (mean (SD)) | 41.03 (15.2) |
| BMI (mean (SD)) | 26.52 (6.2) |
| Biologic administered (%) |  |
| Adalimumab | 5 (5.4) |
| Infliximab | 40 (43.5) |
| Golimumab | 1 (1.1) |
| Ustekinumab | 11 (12) |
| Vedolizumab | 28 (30.4) |
| Tofacitinib | 7 (7.6) |
| Observations by time in week (%) |  |
| Week 0 / baseline | 92 |
| Week 2 | 3 |
| Week 14 | 69 |
| Week 26 | 3 |
| Disease activity (%) / at baseline (%) |  |
| Active disease (eMayo) | 115 (69.3) / 81 (88) |
| Active disease (PRO2) | 122 (73.5) / 82 (90.1) |
| Active disease (Nancy) | 126 (78.8) / 80 (89.9) |
| Active disease (CRP) | 76 (46.1) / 48 (52.8) |
| Remission status week 14 (%) at baseline |  |
| Remission (eMayo) | 39 (58.2) |
| Remission (PRO2) | 34 (49.3) |

**Supplementary Table 3.**
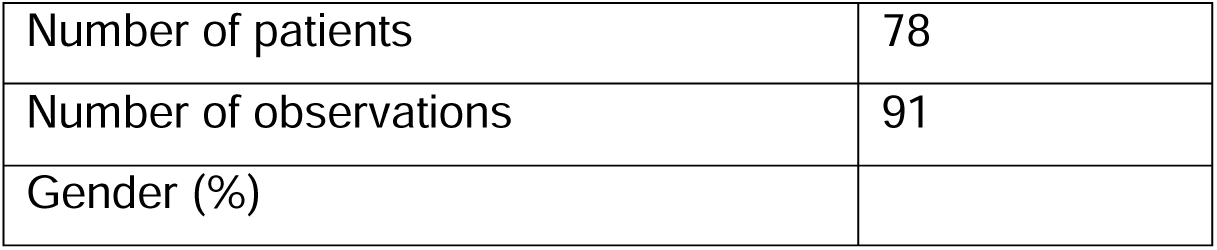
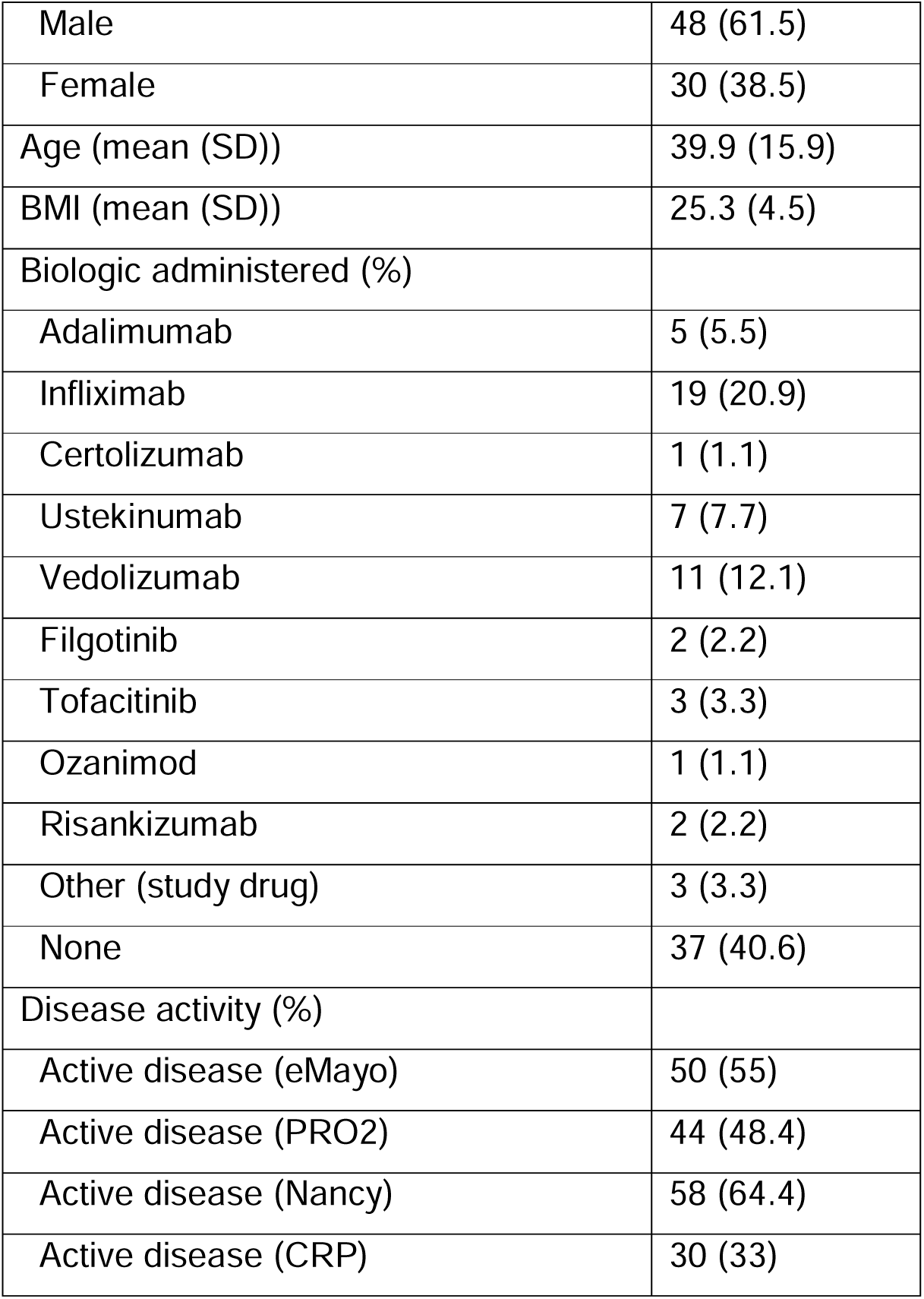
Cohort 2b: Kiel cohort, cross-sectional UC discovery cohort.

|  |  |
| --- | --- |
| Number of patients | 78 |
| Number of observations | 91 |
| Gender (%) |  |
| Male | 48 (61.5) |
| Female | 30 (38.5) |
| Age (mean (SD)) | 39.9 (15.9) |
| BMI (mean (SD)) | 25.3 (4.5) |
| Biologic administered (%) |  |
| Adalimumab | 5 (5.5) |
| Infliximab | 19 (20.9) |
| Certolizumab | 1 (1.1) |
| Ustekinumab | 7 (7.7) |
| Vedolizumab | 11 (12.1) |
| Filgotinib | 2 (2.2) |
| Tofacitinib | 3 (3.3) |
| Ozanimod | 1 (1.1) |
| Risankizumab | 2 (2.2) |
| Other (study drug) | 3 (3.3) |
| None | 37 (40.6) |
| Disease activity (%) |  |
| Active disease (eMayo) | 50 (55) |
| Active disease (PRO2) | 44 (48.4) |
| Active disease (Nancy) | 58 (64.4) |
| Active disease (CRP) | 30 (33) |

**Supplementary Table 4.** Cohort 3: Leuven cohort, longitudinal UC validation cohort.

|  |  |
| --- | --- |
| Number of patients | 52 |
| Number of observations | 103 |
| Gender (%) |  |
| Male | 24 (43.6) |
| Female | 31 (56.4) |
| Age at baseline (mean (SD)) | 44 (15.5) |
| Biologic administered (%) |  |
| Adalimumab | 4 (8) |
| Infliximab | 17 (34) |
| Golimumab | 1 (2) |
| Vedolizumab | 28 (56) |
| Observations by time in week (%) |  |
| Week 0 / baseline | 52 |
| Week 14 | 50 |
| Week 24 | 1 |
| Disease activity (%) / at baseline (%) |  |
| Active disease (eMayo) | 77 (74.8) / 50 (96.2) |
| Active disease (PRO2) | 66 (66) / 45 (90) |
| Active disease (CRP) | 36 (36) / 20 (40) |
| Remission status week 14 (%) at baseline |  |
| Remission (eMayo) | 42 (46.2) |
| Remission (PRO2) | 52 (58.4) |

**Supplementary Table 5.**
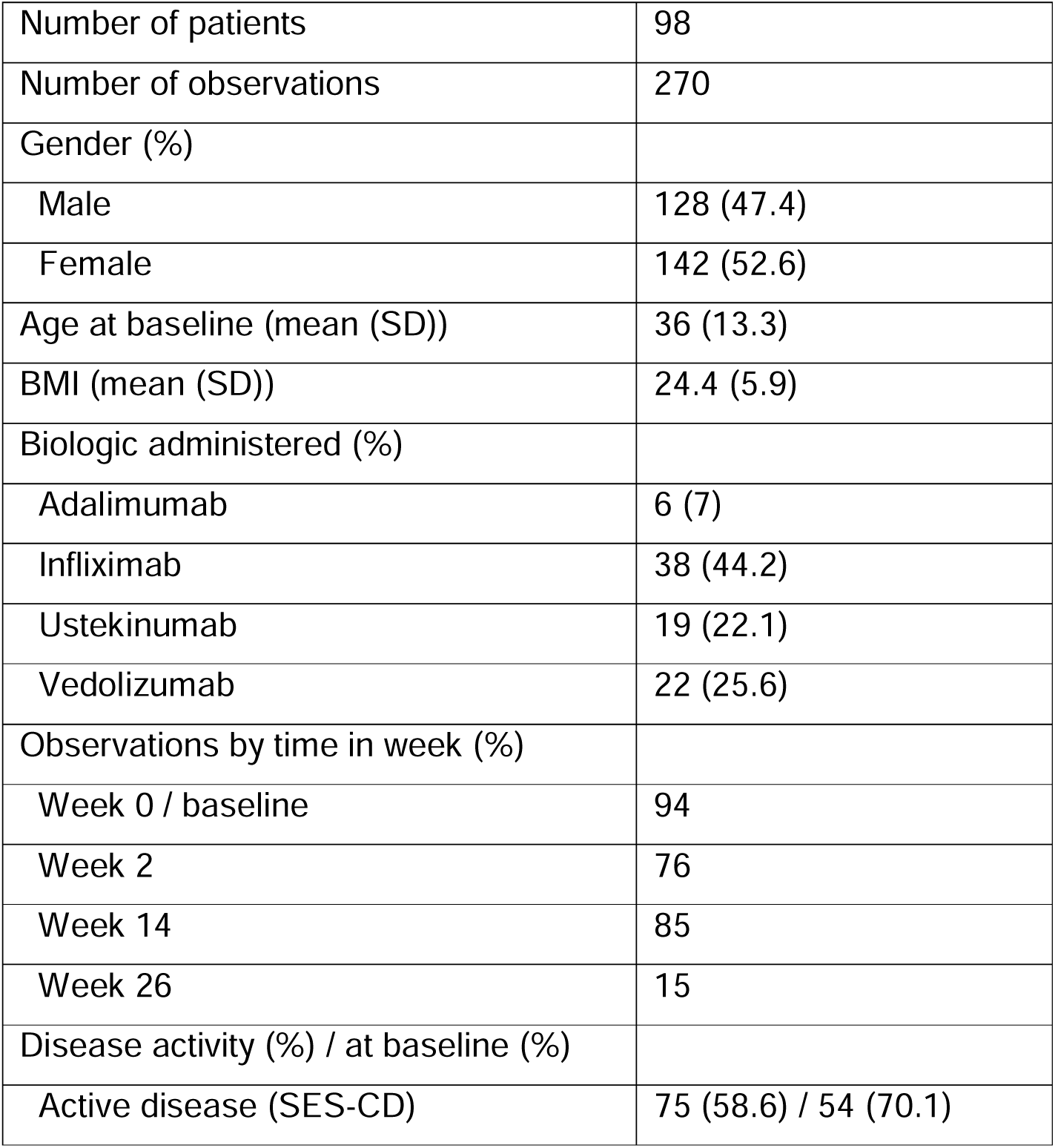
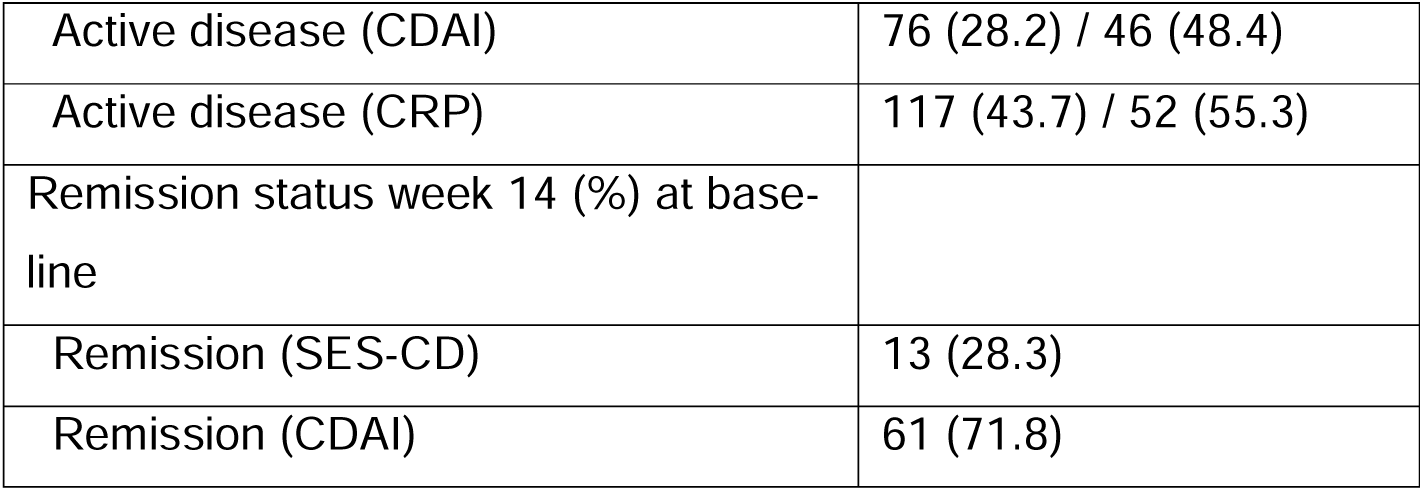
Cohort 4: Kiel cohort, longitudinal CD cohort.

|  |  |
| --- | --- |
| Number of patients | 98 |
| Number of observations | 270 |
| Gender (%) |  |
| Male | 128 (47.4) |
| Female | 142 (52.6) |
| Age at baseline (mean (SD)) | 36 (13.3) |
| BMI (mean (SD)) | 24.4 (5.9) |
| Biologic administered (%) |  |
| Adalimumab | 6 (7) |
| Infliximab | 38 (44.2) |
| Ustekinumab | 19 (22.1) |
| Vedolizumab | 22 (25.6) |
| Observations by time in week (%) |  |
| Week 0 / baseline | 94 |
| Week 2 | 76 |
| Week 14 | 85 |
| Week 26 | 15 |
| Disease activity (%) / at baseline (%) |  |
| Active disease (SES-CD) | 75 (58.6) / 54 (70.1) |
| Active disease (CDAI) | 76 (28.2) / 46 (48.4) |
| Active disease (CRP) | 117 (43.7) / 52 (55.3) |
| Remission status week 14 (%) at base-line |  |
| Remission (SES-CD) | 13 (28.3) |
| Remission (CDAI) | 61 (71.8) |

## References

1 Schreiber S, Rosenstiel P, Albrecht M, et al. Genetics of Crohn disease, an archetypal inflammatory barrier disease. Nat Rev Genet. 2005;6:376–88. doi: 10.1038/nrg1607

2 Khor B, Gardet A, Xavier RJ. Genetics and pathogenesis of inflammatory bowel disease. Nature. 2011;474:307–17. doi: 10.1038/nature10209

3 Piovani D, Danese S, Peyrin-Biroulet L, et al. Environmental Risk Factors for Inflammatory Bowel Diseases: An Umbrella Review of Meta-analyses. Gastroenterology. 2019;157:647–659.e4. doi: 10.1053/j.gastro.2019.04.016

4 Franzosa EA, Sirota-Madi A, Avila-Pacheco J, et al. Gut microbiome structure and metabolic activity in inflammatory bowel disease. Nat Microbiol. 2019;4:293–305. doi: 10.1038/s41564-018-0306-4

5 Neurath MF. Current and emerging therapeutic targets for IBD. Nat Rev Gastroenterol Hepatol. 2017;14:269–78. doi: 10.1038/nrgastro.2016.208

6 Raine T, Danese S. Breaking Through the Therapeutic Ceiling: What Will It Take? Gastroenterology. 2022;162:1507–11. doi: 10.1053/j.gastro.2021.09.078

7 Vermeire S, Van Assche G, Rutgeerts P. Laboratory markers in IBD: useful, magic, or unnecessary toys? Gut. 2006;55:426–31. doi: 10.1136/gut.2005.069476

8 Schreiber S, Danese S, Dignass A, et al. Defining Comprehensive Disease Control for Use as a Treatment Target for Ulcerative Colitis in Clinical Practice: International Delphi Consensus Recommendations. J Crohns Colitis. 2024;18:91–105. doi: 10.1093/ecco-jcc/jjad130

9 Schreiber S, Aden K, Tran F, et al. Rise of precision medicine: can it deliver on its promise in IBD? Gut. 2025;gutjnl-2023-330000. doi: 10.1136/gutjnl-2023-330000

10 Scoville EA, Allaman MM, Brown CT, et al. Alterations in Lipid, Amino Acid, and Energy Metabolism Distinguish Crohn’s Disease from Ulcerative Colitis and Control Subjects by Serum Metabolomic Profiling. Metabolomics. 2018;14:17. doi: 10.1007/s11306-017-1311-y

11 Gallagher K, Catesson A, Griffin JL, et al. Metabolomic Analysis in Inflammatory Bowel Disease: A Systematic Review. J Crohns Colitis. 2021;15:813–26. doi: 10.1093/ecco-jcc/jjaa227

12 Harris DMM, Szymczak S, Schuchardt S, et al. Tryptophan degradation as a systems phenomenon in inflammation - an analysis across 13 chronic inflammatory diseases. EBioMedicine. 2024;102:105056. doi: 10.1016/j.ebiom.2024.105056

13 Starke S, Harris DMM, Zimmermann J, et al. Amino acid auxotrophies in human gut bacteria are linked to higher microbiome diversity and long-term stability. ISME J. 2023;17:2370–80. doi: 10.1038/s41396-023-01537-3

14 Smilde AK, van der Werf MJ, Bijlsma S, et al. Fusion of Mass Spectrometry-Based Metabolomics Data. Anal Chem. 2005;77:6729–36. doi: 10.1021/ac051080y

15 Taubenheim J, Kadibalban AS, Zimmermann J, et al. Metabolic modeling reveals a multi-level deregulation of host-microbiome metabolic networks in IBD. Nat Commun. 2025;16:5120. doi: 10.1038/s41467-025-60233-2

16 Son DO, Satsu H, Shimizu M. Histidine inhibits oxidative stress- and TNF-alphainduced interleukin-8 secretion in intestinal epithelial cells. FEBS Lett. 2005;579:4671–7. doi: 10.1016/j.febslet.2005.07.038

17 Martínez Y, Li X, Liu G, et al. The role of methionine on metabolism, oxidative stress, and diseases. Amino Acids. 2017;49:2091–8. doi: 10.1007/s00726-017-2494-2

18 Ooi M, Nishiumi S, Yoshie T, et al. GC/MS-based profiling of amino acids and TCA cycle-related molecules in ulcerative colitis. Inflammation Research. 2011;60:831–40. doi: 10.1007/s00011-011-0340-7

19 Hisamatsu T, Okamoto S, Hashimoto M, et al. Novel, objective, multivariate biomarkers composed of plasma amino acid profiles for the diagnosis and assessment of inflammatory bowel disease. PLoS One. 2012;7:e31131. doi: 10.1371/journal.pone.0031131

20 Sofia MA, Ciorba MA, Meckel K, et al. Tryptophan Metabolism through the Kynurenine Pathway is Associated with Endoscopic Inflammation in Ulcerative Colitis. Inflamm Bowel Dis. 2018;24:1471–80. doi: 10.1093/ibd/izy103

21 Rüsing S, Welz L, Pfitzer C, et al. Decreased Serum Tryptophan and Severe Ulcerative Damage of Colon Mucosa Identify Inflammatory Bowel Disease Patients With High Risk of Cytomegalovirus Colitis. Clin Transl Gastroenterol. 2024;15:e00731. doi: 10.14309/ctg.0000000000000731

22 Nikolaus S, Schulte B, Al-Massad N, et al. Increased Tryptophan Metabolism Is Associated With Activity of Inflammatory Bowel Diseases. Gastroenterology. 2017;153:1504–1516.e2. doi: 10.1053/j.gastro.2017.08.028

23 Li Q, de Oliveira Formiga R, Puchois V, et al. Microbial metabolite indole-3-propionic acid drives mitochondrial respiration in CD4+ T cells to confer protection against intestinal inflammation. Nat Metab. Published Online First: 21 October 2025. doi: 10.1038/s42255-025-01396-6

24 Hisamatsu T, Ono N, Imaizumi A, et al. Decreased Plasma Histidine Level Predicts Risk of Relapse in Patients with Ulcerative Colitis in Remission. PLoS One. 2015;10:e0140716. doi: 10.1371/journal.pone.0140716

25 Andou A, Hisamatsu T, Okamoto S, et al. Dietary Histidine Ameliorates Murine Colitis by Inhibition of Proinflammatory Cytokine Production From Macrophages. Gastroenterology. 2009;136:564–574.e2. doi: 10.1053/j.gastro.2008.09.062

26 Aldars-García L, Gil-Redondo R, Embade N, et al. Serum and Urine Metabolomic Profiling of Newly Diagnosed Treatment-Naïve Inflammatory Bowel Disease Patients. Inflamm Bowel Dis. 2024;30:167–82. doi: 10.1093/ibd/izad154

27 Dawiskiba T, Deja S, Mulak A, et al. Serum and urine metabolomic fingerprinting in diagnostics of inflammatory bowel diseases. World J Gastroenterol. 2014;20:163–74. doi: 10.3748/wjg.v20.i1.163

28 Watanabe M, Suliman ME, Qureshi AR, et al. Consequences of low plasma histidine in chronic kidney disease patients: associations with inflammation, oxidative stress, and mortality. Am J Clin Nutr. 2008;87:1860–6. doi: 10.1093/ajcn/87.6.1860

29 Gerber DA. Low free serum histidine concentration in rheumatoid arthritis. A measure of disease activity. J Clin Invest. 1975;55:1164–73. doi: 10.1172/JCI108033

30 Santoru ML, Piras C, Murgia A, et al. Cross sectional evaluation of the gutmicrobiome metabolome axis in an Italian cohort of IBD patients. Sci Rep. 2017;7:9523. doi: 10.1038/s41598-017-10034-5

31 Bourgonje AR, Ibing S, Argmann C, et al. DOP33 Distinct perturbances in metabolic pathways associate with disease progression in patients with Inflammatory Bowel Disease. J Crohns Colitis. 2024;18:i131–3. doi: 10.1093/ecco-jcc/jjad212.0073

32 Elger T, Huss M, Liebisch G, et al. Elevated long-to-very-long-chain ceramide ratio correlates with disease severity in inflammatory bowel disease and primary sclerosing cholangitis. Sci Rep. 2025;15:20294. doi: 10.1038/s41598-025-07308-8

33 Salihovic S, Nyström N, Mathisen CB-W, et al. Identification and validation of a bloodbased diagnostic lipidomic signature of pediatric inflammatory bowel disease. Nat Commun. 2024;15:4567. doi: 10.1038/s41467-024-48763-7

34 Mosli MH, Zou G, Garg SK, et al. C-Reactive Protein, Fecal Calprotectin, and Stool Lactoferrin for Detection of Endoscopic Activity in Symptomatic Inflammatory Bowel Disease Patients: A Systematic Review and Meta-Analysis. American Journal of Gastroenterology. 2015;110:802–19. doi: 10.1038/ajg.2015.120

35 Nguyen NH, Picetti D, Dulai PS, et al. Machine Learning-based Prediction Models for Diagnosis and Prognosis in Inflammatory Bowel Diseases: A Systematic Review. J Crohns Colitis. 2022;16:398–413. doi: 10.1093/ecco-jcc/jjab155

36 Prins FM, Hidding IJ, Klaassen MAY, et al. Limited predictive value of the gut microbiome and metabolome for response to biological therapy in inflammatory bowel disease. Gut Microbes. 2024;16:2391505. doi: 10.1080/19490976.2024.2391505

37 Sazonovs A, Schut K, Plevris N, et al. OP19 Preand Post-diagnostic Metabolomic Biomarker Profiling of Over 700,000 Individuals in Three National Biobanks Enables Prediction of Inflammatory Bowel Disease Onset and Complications. J Crohns Colitis. 2025;19:i38–40. doi: 10.1093/ecco-jcc/jjae190.0019

