## Supplementary figures and images for "A distinct serum metabolic signature outperforms C-reactive protein as a non-invasive marker for monitoring disease activity in Inflammatory Bowel Disease"

### Supplementary figures

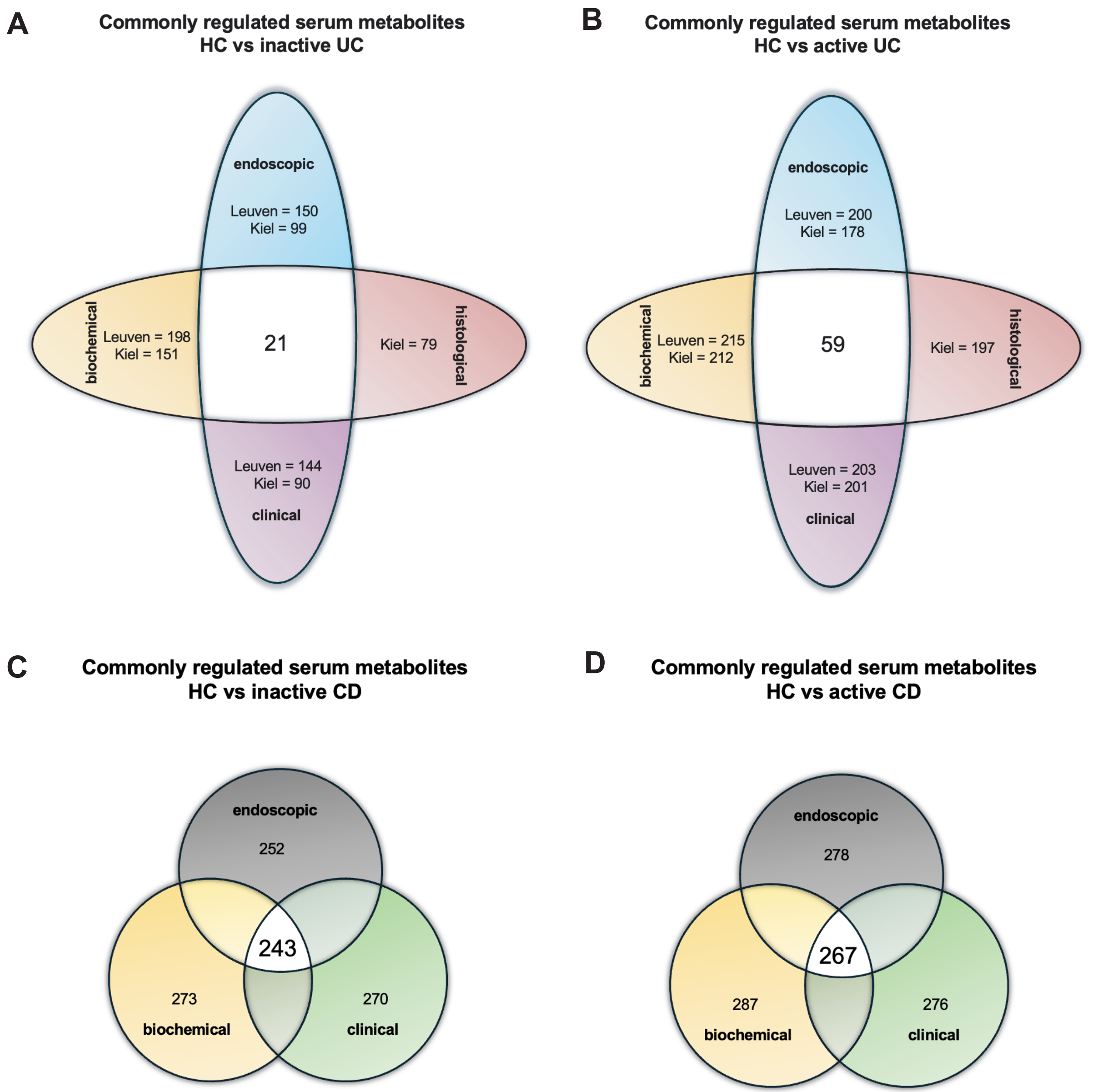

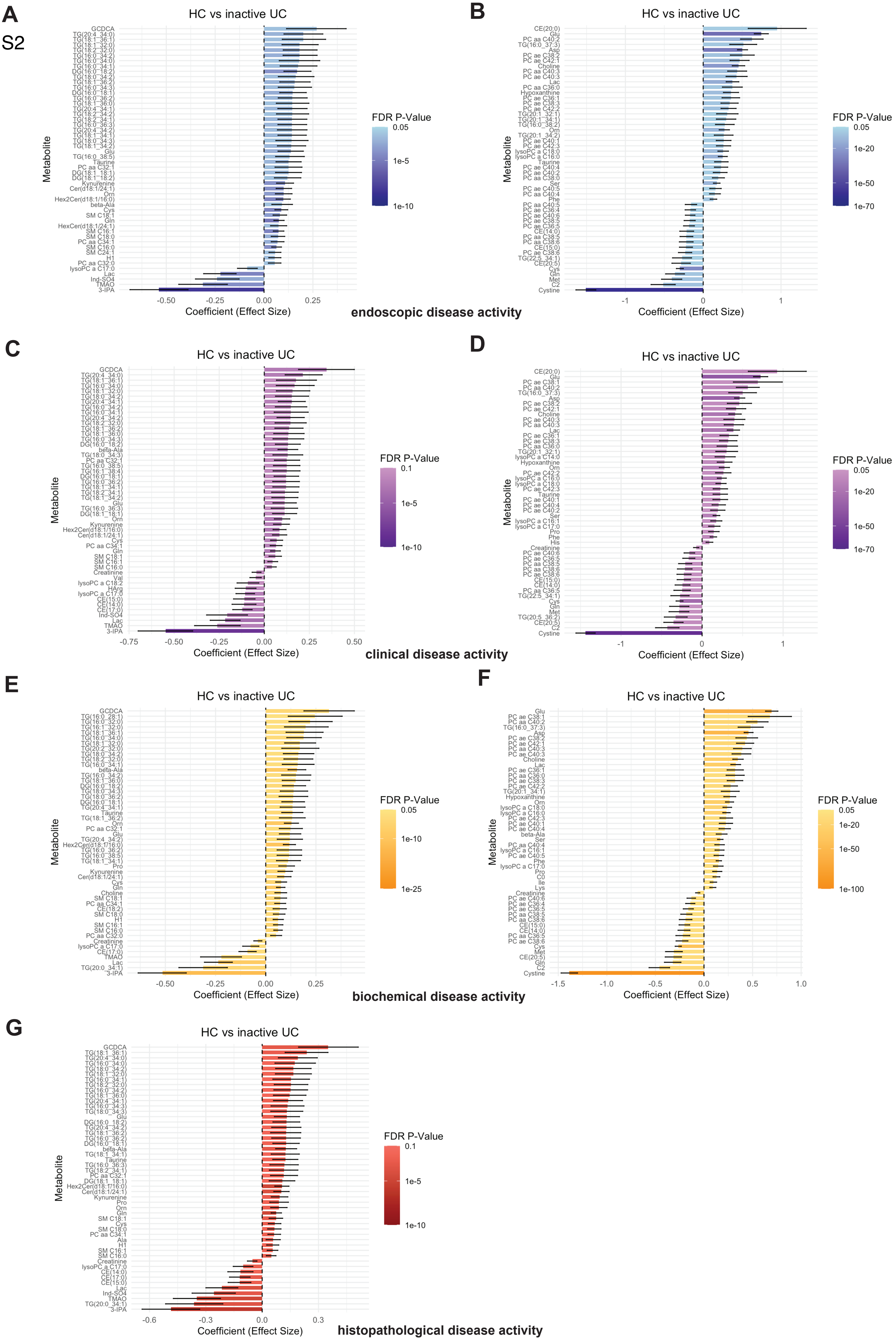

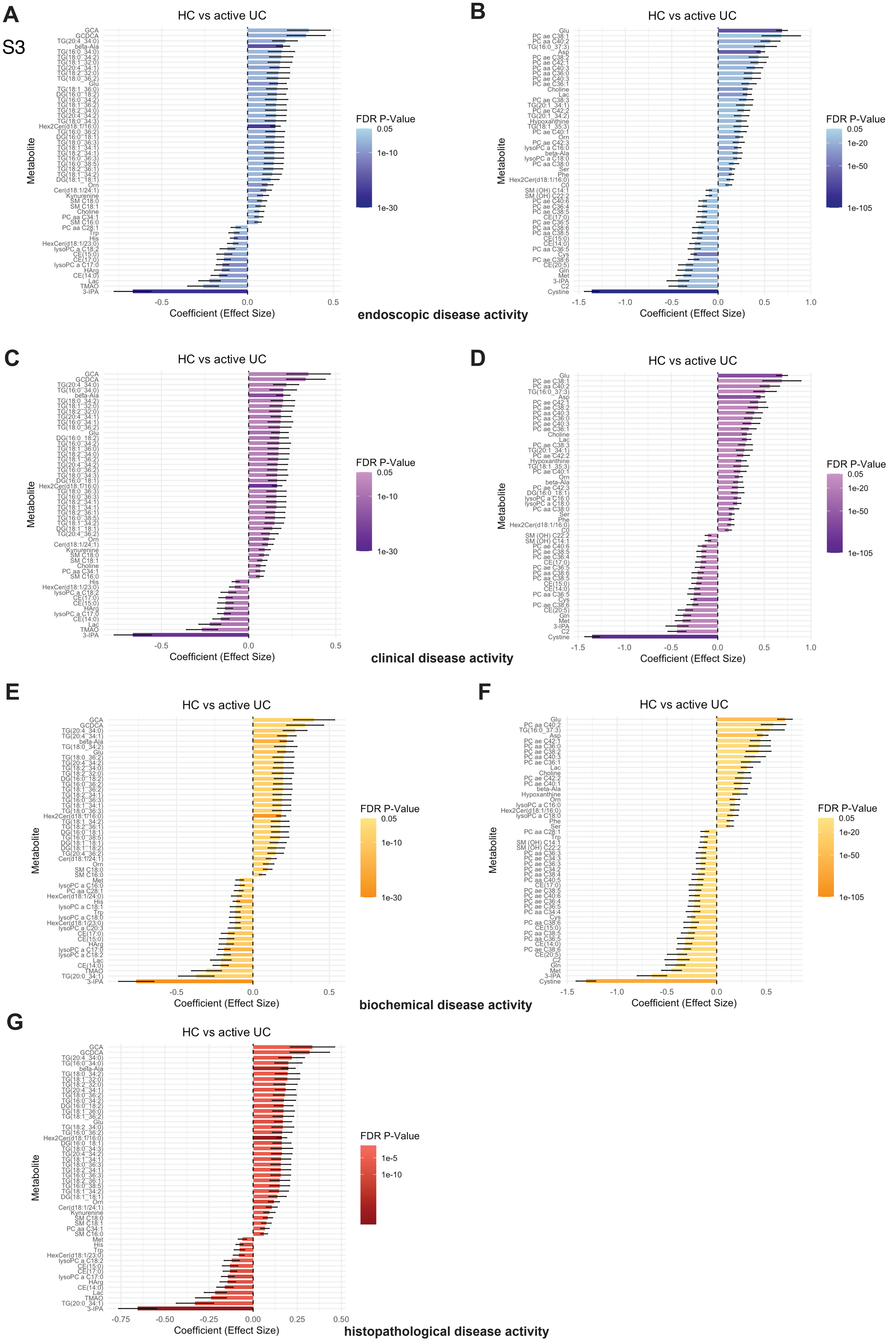

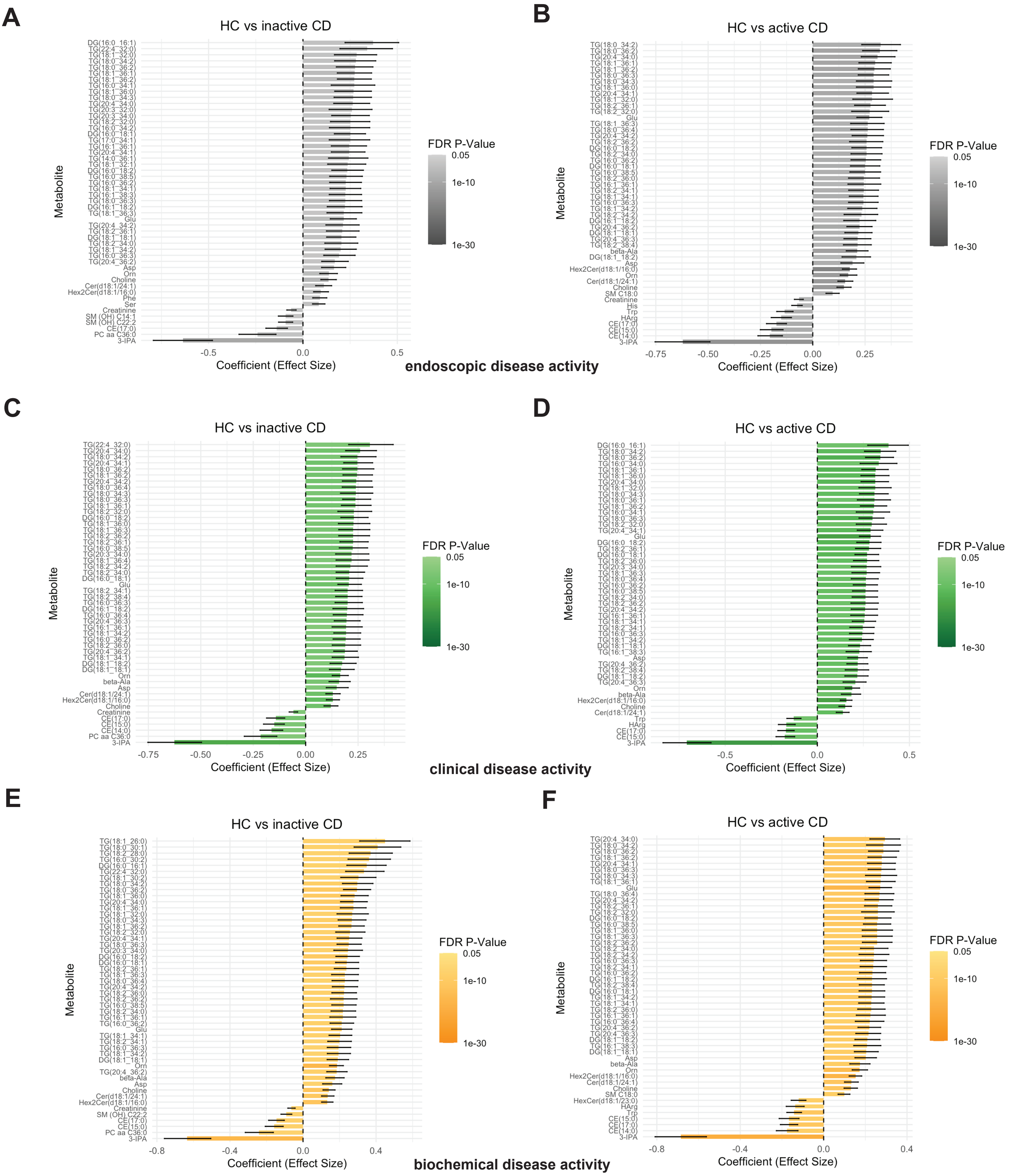

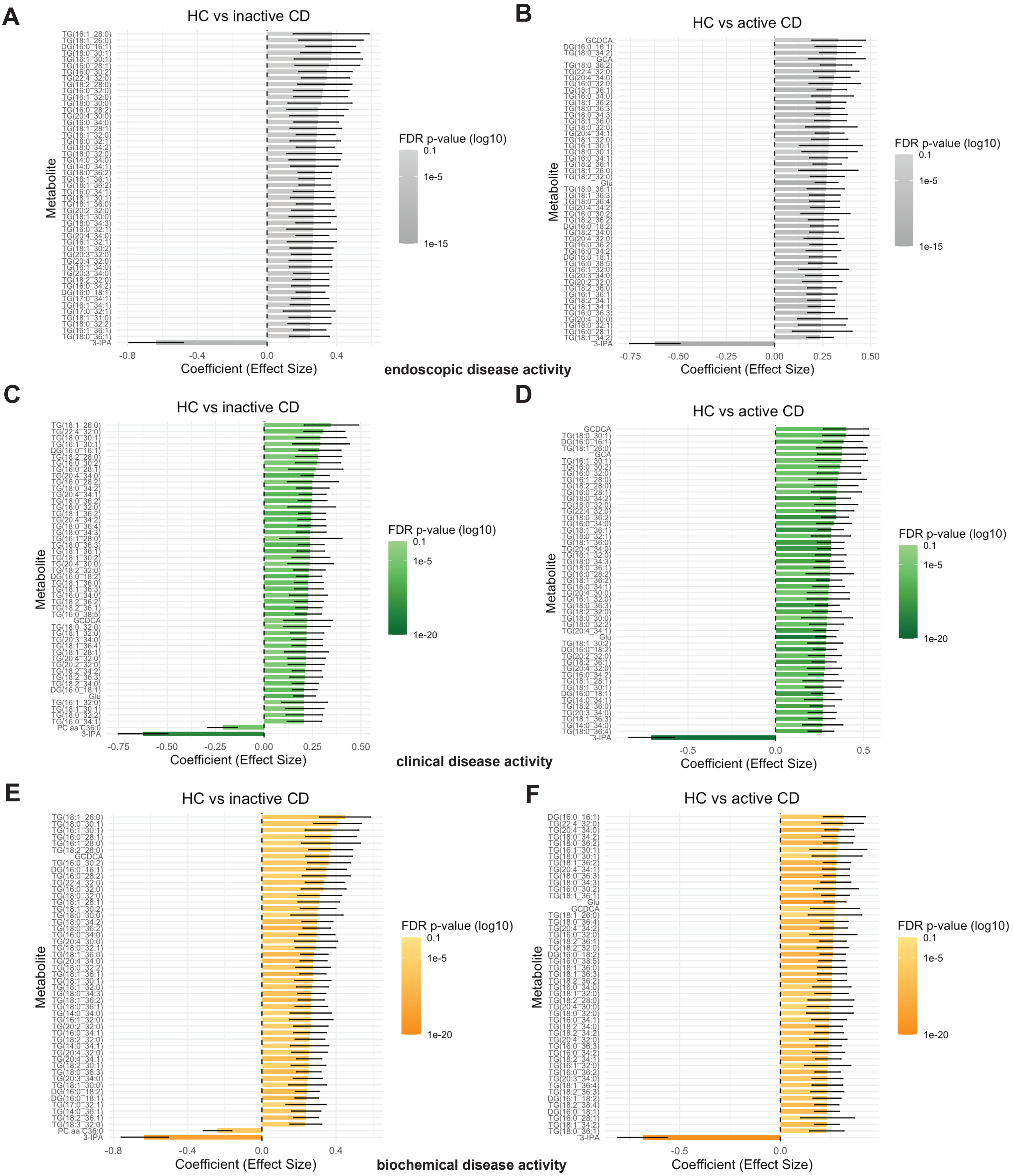

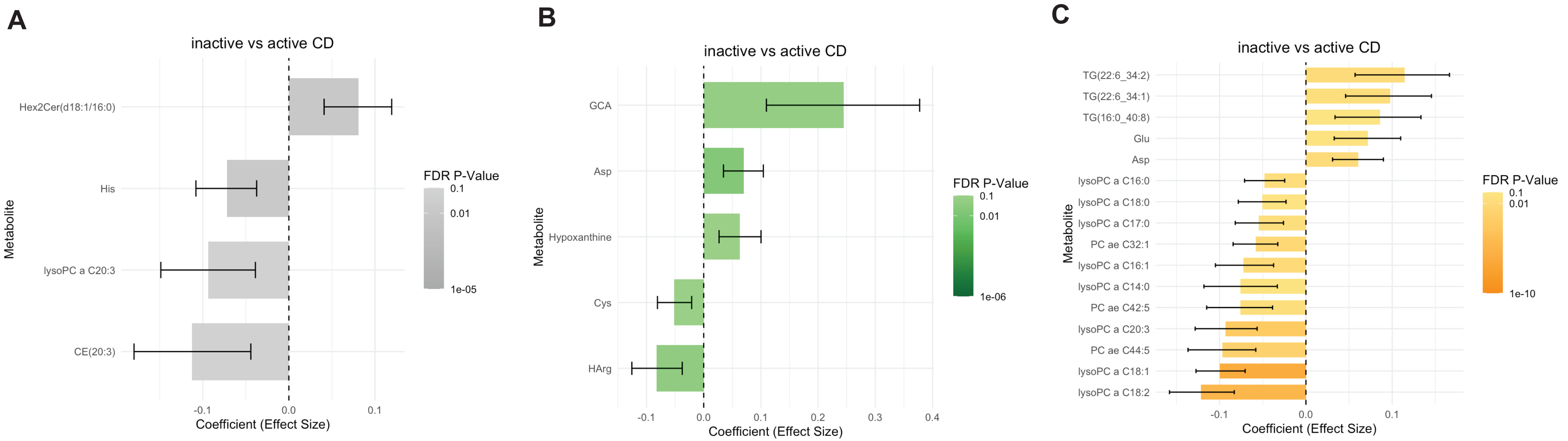

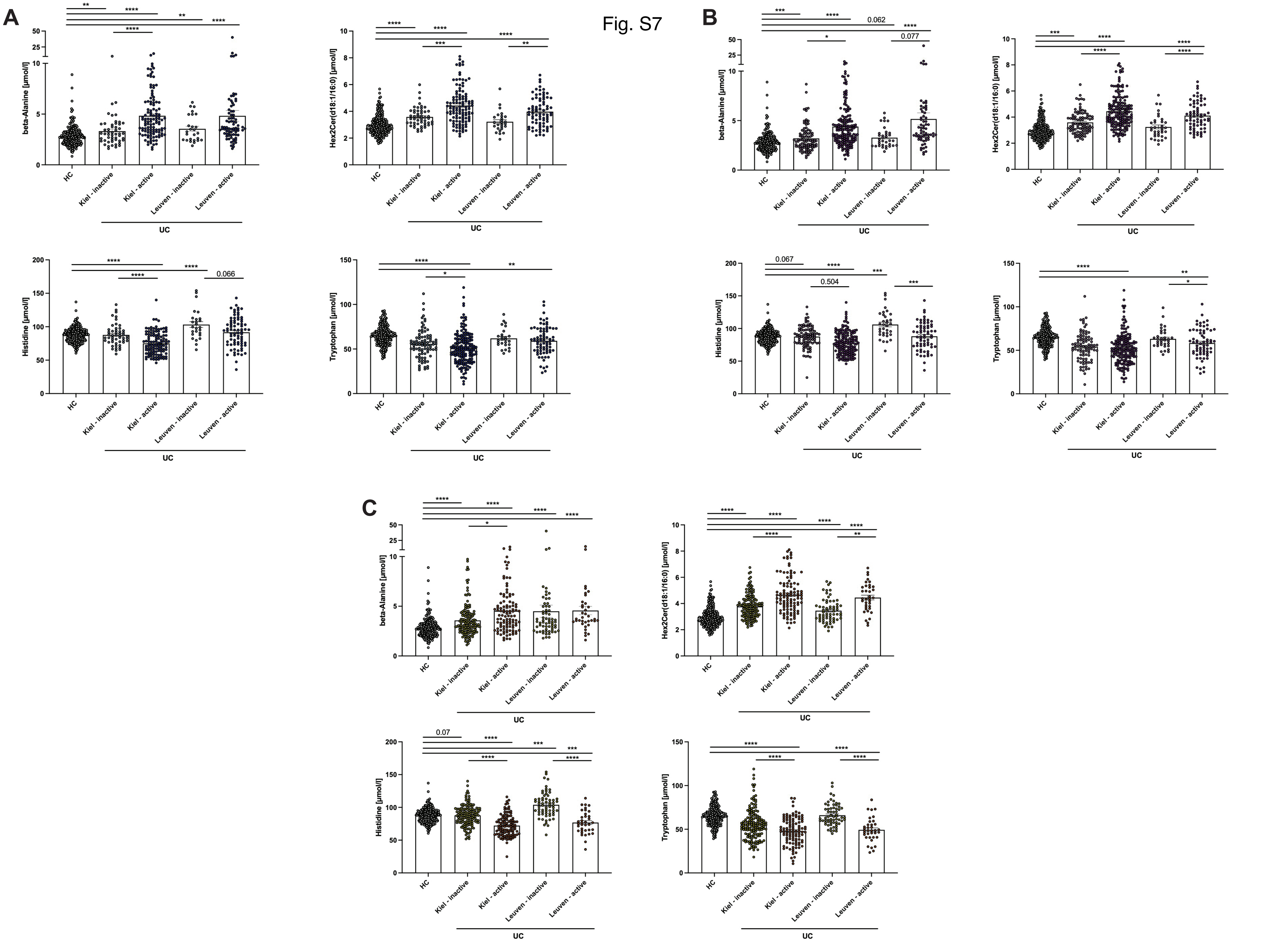

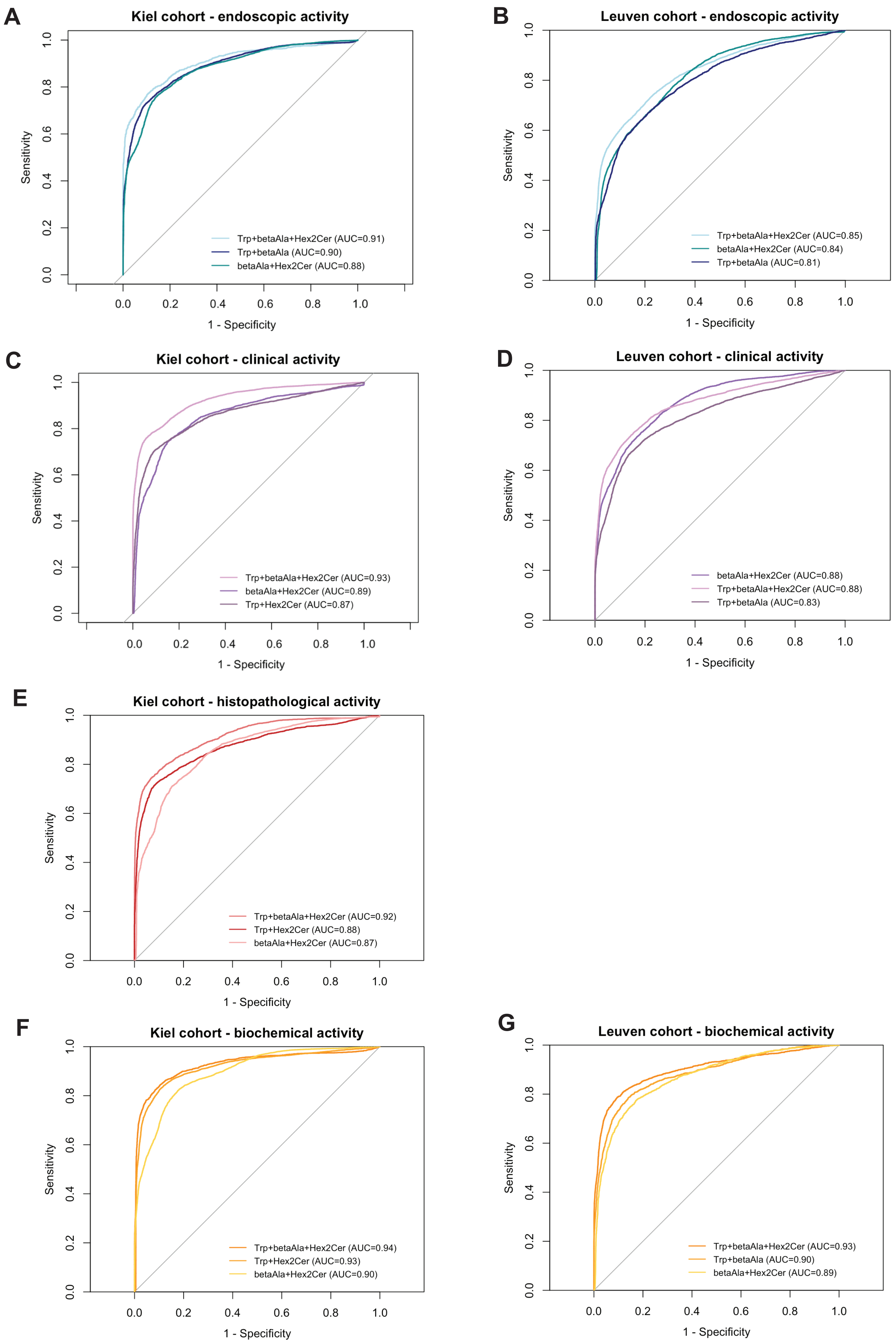

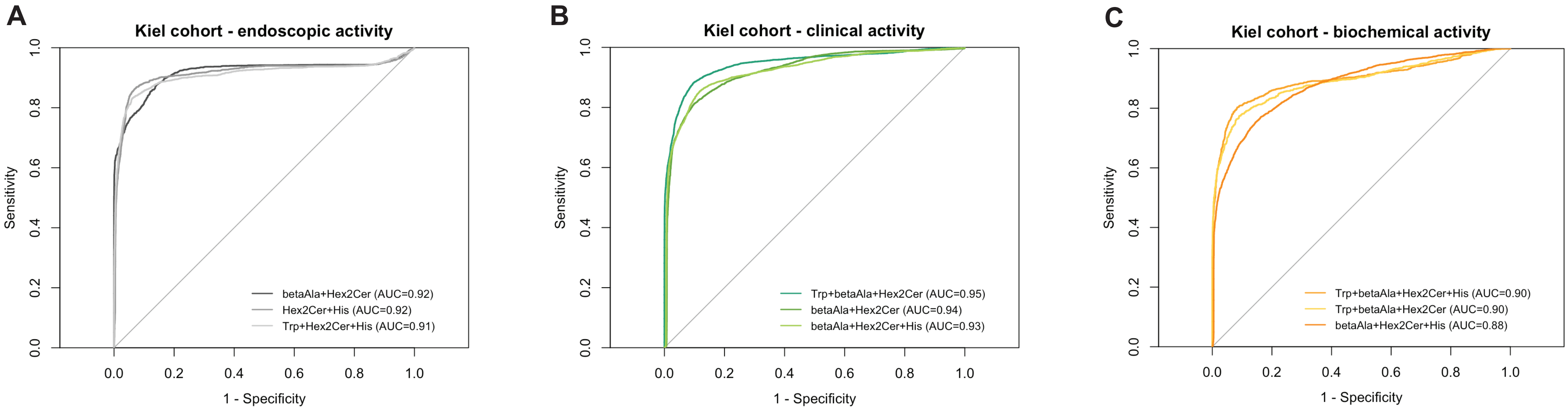

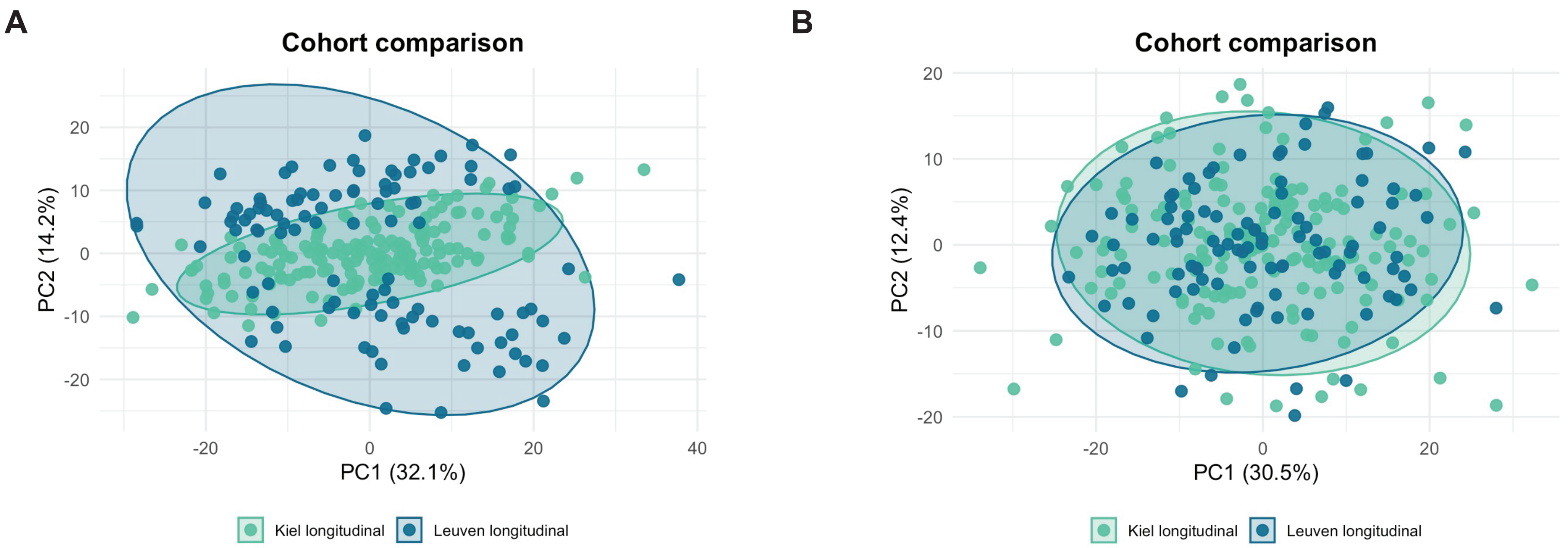
